# Intensity of public health and social measures are associated with effectiveness of SARS-CoV-2 vaccine in test-negative study

**DOI:** 10.1101/2025.05.08.25327221

**Authors:** Tim K. Tsang, Sheena G. Sullivan, Xiaotong Huang, Can Wang, Liping Peng, Bingyi Yang, Benjamin J. Cowling

## Abstract

The intensity and duration of exposure can influence vaccine effectiveness (VE). For “leaky” vaccines such as SARS-CoV-2 vaccines, which reduce but do not entirely prevent infections, repeated or prolonged exposures may increase breakthrough infection likelihood. To test this hypothesis, we conducted a systematic review and meta-analysis of 76 test-negative design studies reporting VE against SARS-CoV-2 infection or severe disease. Exposure intensity was approximated using Oxford COVID-19 Government Response Tracker indices: Stringency Index (SI), Containment and Health Index (CHI), and Government Response Index (GRI). Based on 1,419 VE estimates, pooled VE against infection was significantly higher in settings with higher index values (lower exposure intensity): 82% (95% CI: 80-83%) in high-SI settings versus 39% (95% CI: 35-43%) in low-SI settings. Similar patterns appeared for other indices and severe disease outcomes. These associations persisted in meta-regression models adjusting for viral variant, vaccine type, time since vaccination, prior infection status, and enrollment criteria. Correlation analyses showed moderate-to-strong positive correlations between VE estimates and exposure indices (Spearman’s correlation: 0.50-0.62). These findings establish exposure intensity as a critical effect modifier of SARS-CoV-2 VE, demonstrating the leaky nature of COVID-19 vaccines and explaining heterogeneity in real-world effectiveness estimates. Future VE evaluations and vaccination strategies should account for exposure intensity to ensure accurate, context-specific estimates.

## INTRODUCTION

The COVID-19 pandemic, caused by SARS-CoV-2, has underscored the importance of effective vaccination strategies to reduce the burden of disease and safeguard public health. While vaccines have demonstrated significant efficacy in controlled clinical trials, their effectiveness in real-world conditions often varies due to differences in exposure, study design, and population characteristics. Vaccine effectiveness (VE) against COVID-19 is influenced by the level and duration of exposure to SARS-CoV-2, which is shaped by public health measures and individual behaviors. Observational studies, particularly test-negative design (TND) studies, have been widely used to evaluate VE in real-world settings (1). However, substantial variability in VE against COVID-19 has been reported both between and within populations (2-5). Some of this heterogeneity may be attributable to differences in the vaccines used and study design choices, such as the use of clinical symptoms criteria (6, 7), which we have previously shown can influence VE (8).

VE is also influenced by underlying population susceptibility, which itself may depend on pre-existing population immunity arising from prior infections (9) as well as behavioral differences affecting the amount of contact between potentially infectious and susceptible individuals (10). Directly measuring whether an individual has been exposed to SARS-CoV-2 (sometimes called the exposure-necessity assumption (11)) is challenging due to the dynamic nature of viral transmission and the complexity of population behaviours (12-15). Consequently, VE studies typically assume that vaccinated and unvaccinated individuals have equal risk of infection (16). However, it has previously been shown that vaccines with incomplete or “leaky” protection may demonstrate reduced effectiveness in high-exposure settings, where more frequent or more intense viral encounters could overcome vaccine-induced immunity (17) (18). Conversely, in settings with stringent public health measures, lower exposure levels may allow vaccines to perform more effectively by reducing the likelihood of breakthrough infections. In contrast, “all-or-nothing” vaccines provide complete immunity to some individuals while leaving others entirely susceptible (18, 19). In this model, VE is determined by the proportion of fully immune individuals and remains unaffected by exposure intensity.

Uniquely, during the COVID-19 pandemic several indicators of public health control measures to limit viral transmission were maintained which may make it possible to examine the role of differences in duration and intensity of exposure on COVID-19 VE (20). Such indices systematically quantify public health interventions such as lockdowns, mask mandates, and travel restrictions. Higher values reflect stricter measures and reduced opportunity for exposure, while lower values indicate weaker measures and greater opportunities for prolonged or frequent viral encounters (11, 17). In this study, we use three indices as proxies to investigate the relationship between exposure intensity to SARS-CoV-2 and estimated VE: the Stringency Index (SI); Containment and Health Index (CHI); and Government Response Index (GRI). We hypothesize that higher exposure intensity, indicated by lower index values, are associated with lower VE, reflecting the potential for COVID-19 vaccines to exhibit “leaky” behaviour in high-exposure settings. By examining how these indices act as effect modifiers and confounders, this study provides critical insights into the context-dependent nature of VE and offers guidance for optimizing vaccination strategies in diverse public health environments.

## RESULTS

### Overview

We systematically searched PubMed, Embase, and Web of Science for test-negative design studies reporting vaccine effectiveness (VE) against SARS-CoV-2 infection or severe disease following primary vaccination series. A total of 13,475 studies were identified, of which 5,783 were duplicates. Following title and abstract screening of the remaining articles, 864 studies were selected for full-text review. From these, 76 studies met the inclusion criteria (3, 4, 21-94). (Figure 1; Table S2–S3). Across these studies, 924 VE estimates against infection were extracted from 63 studies, while 495 estimates against severe disease were derived from 49 studies (Table S4). A detailed summary of study characteristics and the distribution of VE estimates across various subgroups, such as prior infection status, enrollment criteria, vaccine types, and circulating virus variants, is provided in Table S5.

**Figure 1.**
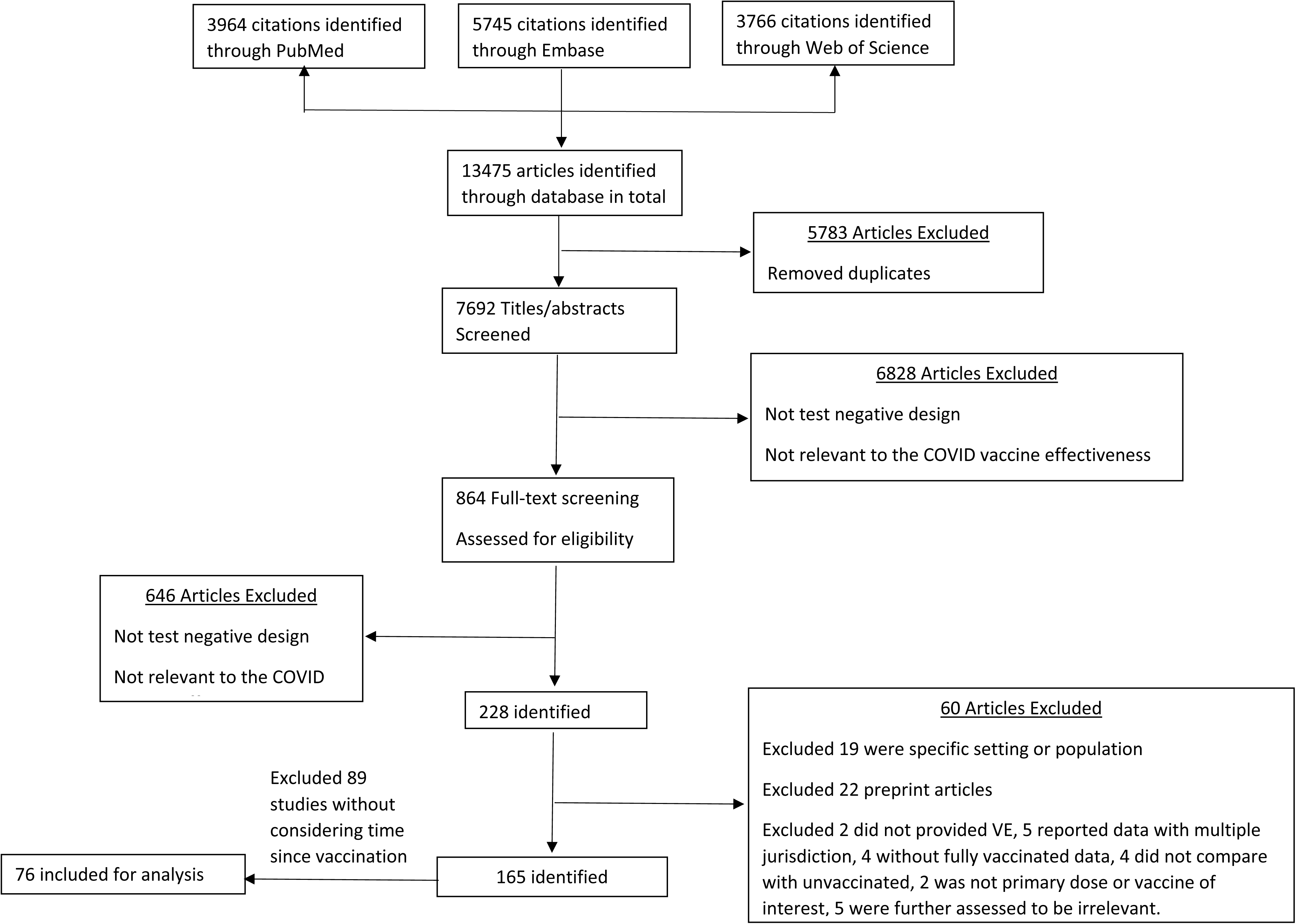
Selection of studies for the systematic review

The primary focus of the study was to examine the relationship between VE and potential exposure intensity. For a leaky vaccine, we hypothesized that more frequent or more intense exposures would be associated with reduced VE (Figure S1). Exposure intensity was approximated using indices from the Oxford COVID-19 Government Response Tracker as proxies for exposure intensity: the Stringency Index (SI), Containment and Health Index (CHI), and Government Response Index (GRI). Higher values of these indices (scale 0-100) reflect stricter public health measures and consequently lower exposure intensity.

We analyzed 1,419 VE estimates from 76 eligible studies, stratified by outcome (infection or severe disease), circulating variant, vaccine type, and time since vaccination. We conducted random-effects meta-analyses to estimate pooled VE across tertiles of each index and performed meta-regression to assess the relationship between indices and VE while adjusting for potential confounders. Correlation analyses using Pearson and Spearman coefficients quantified the association between exposure indices and VE estimates.

Overall, we observed moderate to high positive correlations between VE against infection and severe disease with SI, CHI and GRI, except in the case of the Omicron BA.4/BA.5 subvariant (Figure 2, Table S1). These suggested that lower exposure intensity, proxied by these indices, was associated with higher VE estimates.

**Figure 2.**
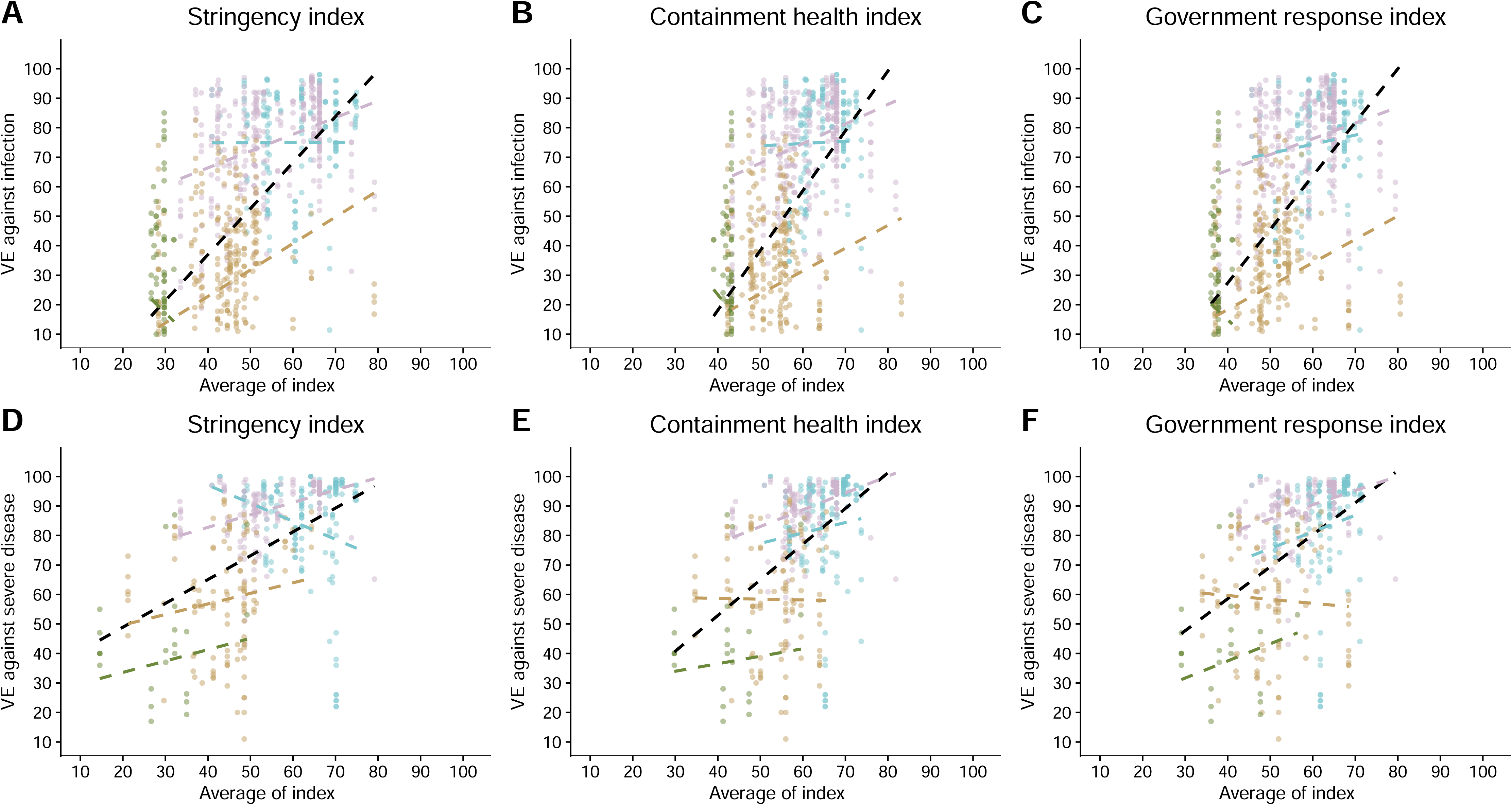
Association between vaccine effectiveness (VE) and public health indices. Scatterplots illustrating the relationship between VE and the Stringency Index (SI), Containment and Health Index (CHI), and Government Response Index (GRI) for different outcomes (infection and severe disease). Correlation coefficients (r and ρ) are shown for the overall dataset and stratified by variant periods (pre-Delta and Delta, late-Delta, Omicron BA.1/BA.2, Omicron BA.4/BA.5). The figures demonstrate positive correlations between VE and higher index values, indicating that stricter public health measures are associated with increased VE.

### Vaccine effectiveness against infection and severe disease

The 924 VE point estimates against infection exhibited considerable heterogeneity, spanning from -38% to 98%, with an I² value of 100% (Figure 3). The 495 VE estimates against severe disease also demonstrated significant variability, with I²=100% and point estimates ranging between -4% and 100% (Figure 3).

**Figure 3.**
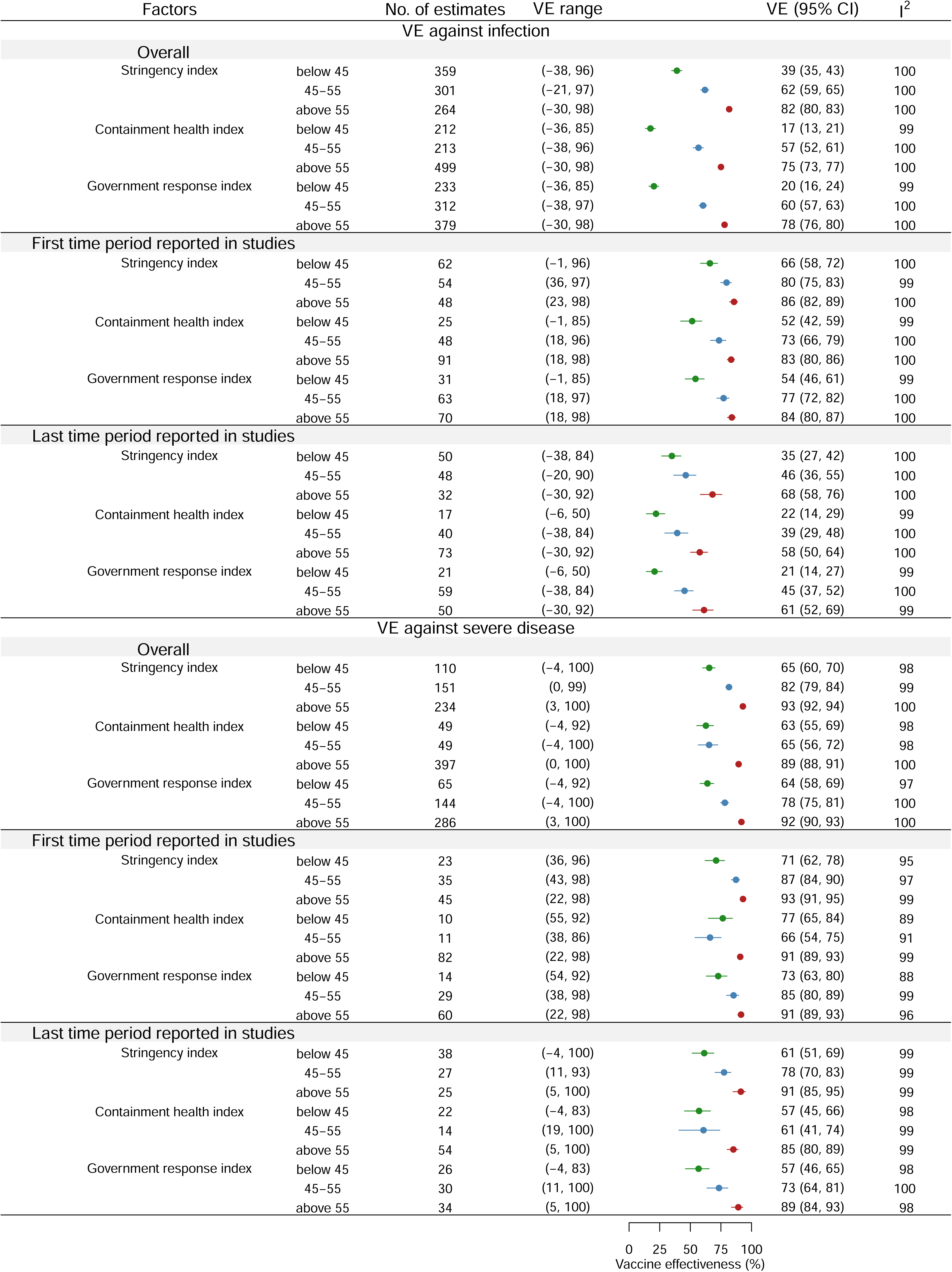
Impact of public health indices on pooled vaccine effectiveness (VE). Meta-analysis results comparing VE across tertiles of the Stringency Index (SI), Containment and Health Index (CHI), and Government Response Index (GRI). The pooled VE is significantly higher in the highest tertile of each index, suggesting that stricter control measures, and consequently lower exposure levels, are associated with improved VE against both infection and severe disease.

In the earliest post-vaccination period (Figure 3), when waning effects were minimal, VE estimates against (n=164) ranged from -0.97% to 97.7%(I²=100%), while severe disease estimates (n=103) ranged from 22% to 98% (I²=99%). In the latest period (Figure 3), as waning effects became pronounced, VE estimates against infection (n=130) varied from -38% to 92% (I²=100%), compared to severe disease estimates (n=90) that ranged from -4% to 100% (I²=100%).

### Impact of intensity of control measure on vaccine effectiveness

Our meta-analysis (Figure 3) demonstrated that higher intensity of control measures and hence lower exposure intensity, measured by SI, CHI and GRI, was associated with higher pooled VE. For VE against infection, high stringency measures resulted in a pooled VE of 82% (95% CI: 80%, 83%), compared to 62% (95% CI: 59%, 65%) for moderate SI and 39% (95% CI: 35%, 43%) for low SI. Similarly, high CHI yielded a pooled VE of 75% (95% CI: 73%, 77%), compared to 57% (95% CI: 52%, 61%) for moderate and 17% (95% CI: 13%, 21%) for low CHI. For GRI, VE against infection was 78% (95% CI: 76%, 80%) for high intensity, 60% (95% CI: 57%, 63%) for moderate intensity, and 20% (95% CI: 16%, 24%) for low intensity.

A similar trend was observed for VE against severe disease. High-intensity SI were associated with a pooled VE of 93% (95% CI: 92%, 94%), compared to 82% (95% CI: 79%, 84%) for moderate SI and 65% (95% CI: 60%, 70%) for low SI. For CHI, VE against severe disease was 89% (95% CI: 88%, 91%) for high intensity, 65% (95% CI: 56%, 72%) for moderate intensity, and 63% (95% CI: 55%, 69%) for low intensity. Similarly, GRI yielded a pooled VE of 92% (95% CI: 90%, 93%) for high intensity, 78% (95% CI: 75%, 81%) for moderate intensity, and 64% (95% CI: 58%, 69%) for low intensity. These patterns were consistent when further analysed for vaccine effectiveness reported in the earliest and the latest period post-vaccination (Figure 3).

In the meta-regression analysis, which adjusted for variables including vaccine type, circulating virus, enrolment criteria, time since vaccination, and prior infection (Table 1, S6), the relative odds ratios (RORs) for infection were 0.84 (95% CI: 0.81, 0.87) for the SI, 0.83 (95% CI: 0.79, 0.88) for the CHI, and 0.85 (95% CI: 0.81, 0.89) for the GRI. These findings suggest that higher control measure intensity and hence lower exposure intensity was associated with improved VE. For instance, in a setting with a baseline VE of 50% against infection, a 10-unit increase in SI, CHI, and GRI would correspond to increases in VE to 58% (95% CI: 56.5%, 59.5%), 58.5% (95% CI: 56%, 60.5%), and 57.5% (95% CI: 55.5%, 59.5%), respectively.

**Table 1.**
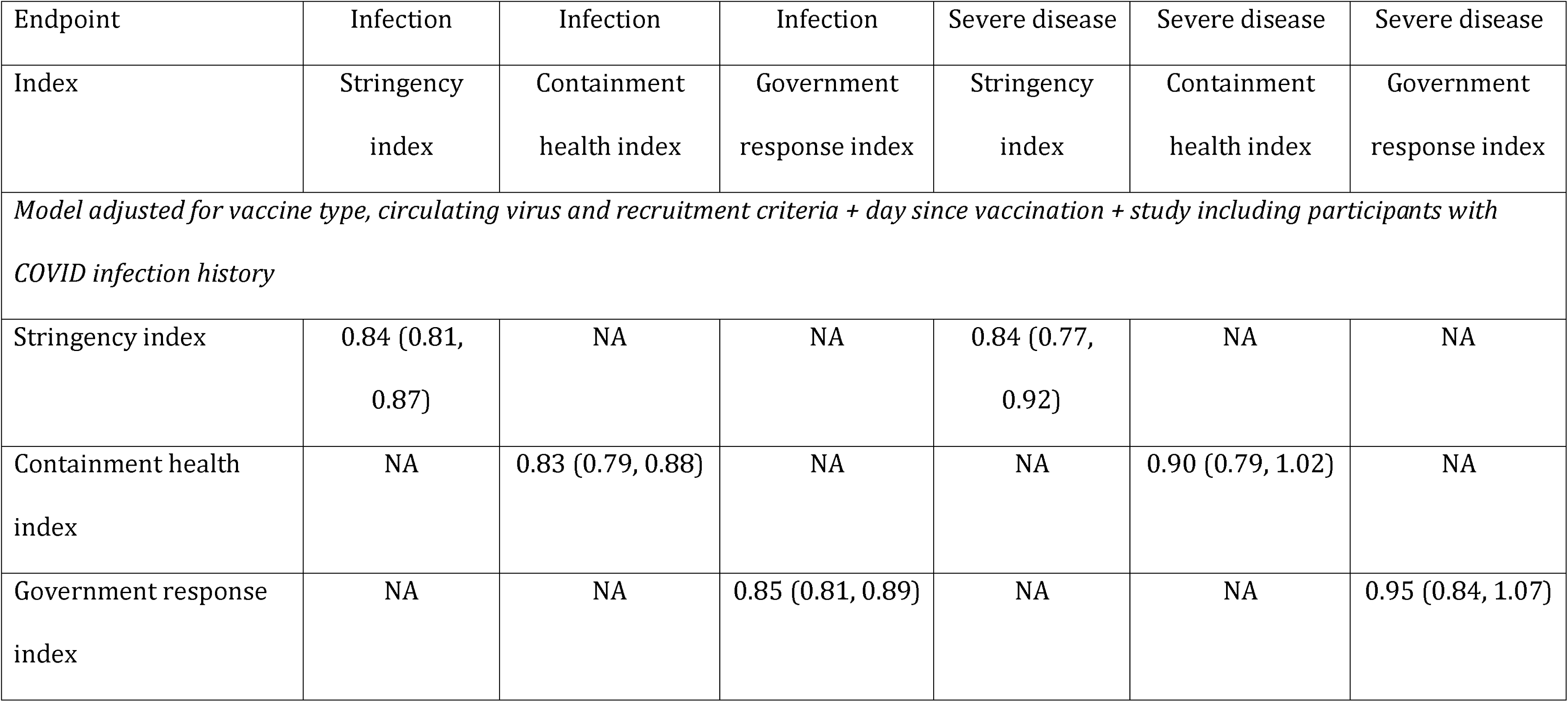
Relationship between government response index and estimates of risk ratios against infection or severe disease.

Similarly, the RORs for severe disease were 0.84 (95% CI: 0.77, 0.92) for SI, but did not reach statistical significance for CHI (0.90; 95% CI: 0.79, 1.02), and for GRI (0.95; 95% CI: 0.84, 1.07). For a baseline VE of 50% against severe disease, a 10-unit increase in SI would result in increases to 58% (95% CI: 54%, 61.5%) respectively. These results remained consistent after excluding estimates from studies identified as having a serious risk of bias (Table S7), and excluding estimates from studies not using a clinical case definition or excluding participants with prior infections (Table S8).

### Impact of type of vaccine, circulating viruses, prior infection and enrolment criteria

Our meta-analysis revealed that pooled VE against infection varied by vaccine type: 63% (95% CI: 61%, 66%) for mRNA vaccines, 66% (95% CI: 61%, 71%) for adenovirus vector vaccines, and 40% (95% CI: 34%, 45%) for inactivated virus vaccines (Figure S3-5). Moreover, VE against infection was markedly lower during Omicron periods: 28% (95% CI: 25–31%) during BA.1/2 and 19% (95% CI: 14–24%) during BA.4/5, compared with 80–81% during the pre-Delta/Delta and late-Delta periods. A similar decline was observed for VE against severe disease, which decreased from 91–93% in the pre-Delta/Delta and late-Delta periods to 61% (95% CI: 57–65%) during BA.1/2 and 33% (95% CI: 27–38%) during BA.4/5. These trends persisted when analyses were stratified by prior infection status and using fixed-effects models (Figure S6). Additionally, studies including participants with prior COVID-19 infection reported higher pooled VE estimates against infection (68% vs. 55%) and severe disease (88% vs. 84%), with a similar pattern observed based on enrollment criteria. Further details of analyses of these factors could be found in Supplementary Note 1.

### Risk of bias

The majority of studies included in the meta-analysis were assessed as having a moderate risk of bias. Specifically, 58 out of 63 studies (92%) analyzing VE against infection and 48 out of 49 studies (98%) evaluating VE against severe disease were categorized as moderate risk (Figure S7–S8). A smaller proportion of studies were judged to have a serious risk of bias: five studies (8%) in the VE against infection analysis and one study (2%) in the VE against severe disease analysis. The primary sources of bias included potential confounding, misclassification of interventions due to self-reported vaccination status, and participant selection bias (Figure S7–S8).

Sensitivity analyses excluding estimates from studies with serious or critical bias (Figure S9; Table S7) or those that excluded participants with prior infections or that did not use a clinical case definition yielded results consistent with the main findings (Figure S10; Table S8). This suggests that the observed impact of control measure intensity on VE estimates remained robust despite the exclusion of higher-risk studies.

## DISCUSSION

In this study, we hypothesized that higher exposure intensity to SARS-CoV-2 would be associated with lower VE, consistent with the related hypothesis that COVID-19 vaccines provide “leaky” protection rather than “all-or-nothing” protection (18, 19). We used three indices (SI, CHI, GRI), to approximate the intensity of public health and social measures (PHSMs) and hence the exposure intensity to SARS-CoV-2. Our findings indicate that higher values of these indices, reflecting stricter public health measures and hence lower exposure intensity, were associated with higher VE estimates. This suggests that regions with stricter control measures may experience reduced exposure intensity, with higher associated VE that would decline when PHSMs are lifted. In addition, vaccine trials conducted in locations and periods with stricter public health measures would likely produce higher estimates of vaccine efficacy than trials or observational studies done in other locations or periods.

The most likely explanation for this relationship is that the exposure intensity to SARS-CoV-2, shaped by PHSMs, acts as an effect modifier of vaccine effectiveness (VE). VE may vary depending on the exposure intensity an individual has to the virus, such as their engagement in high-risk behaviors (95). Another possible explanation is that lower exposure intensity reduces opportunities for the virus to cause an infection in vaccinated individuals, giving their immune systems sufficient time to respond effectively to each encounter and a greater chance to neutralize the virus and prevent infection (96-98), so that the infectious dose is not achieved.

In addition to acting as effect modifiers, SI, CHI, and GRI could also function as confounders in the relationship between COVID-19 vaccination and outcomes. These indices are often correlated with both vaccination strategies and infection risk. For instance, regions with higher levels of PHSMs (higher SI, GRI or CHI) are likely to have more extensive vaccination campaigns, with higher coverage and better access to vaccines. These regions often experienced lower levels of virus circulation due to the stringent measures in place, independent of reduced exposure intensity associated with interventions such as mask wearing that reduces the infection risk after exposures (99, 100) or travel restrictions that prevent virus introduction to a region (101-104).

The relationship between VE and exposure intensity underscores the need to consider PHSMs when interpreting VE data. An important implication of our findings is that cross-country comparisons of VE may be inherently limited due to differing levels of PHSMs. Our analysis indicates that regions with stringent control measures tended to report higher VE, likely reflecting reduced exposure intensity rather than intrinsic differences in vaccine performance. Consequently, for policy makers, it is critical to contextualize VE estimates by considering data from settings with comparable levels of restrictions. This targeted approach not only ensures a more accurate interpretation of VE but also provides valuable insights for guiding the implementation or relaxation of restrictions. These results also support previous assertions that COVID-19 vaccines are leaky (18, 19). This may impact interpretation of VE estimates, For example, depletion of susceptibles bias is a concern for leaky vaccines but not all-or-nothing vaccines, and should therefore be taken into consideration in VE estimation (105). Moreover, the optimal vaccine target group could change depending on whether a vaccine affords all-or-nothing or leaky protection (106) even when the VE is the same.

It is important to recognize that these exposure indices are proxies for the complex reality of exposure intensity. Not all of the heterogeneity in VE can be explained by differences in exposure intensity. We considered additional sources of variation, including vaccine type, circulating virus variant, enrolment criteria, prior infection status, and time since vaccination. Some differences in study design, population demographics, behavioural factors also play critical roles. Therefore, while our findings underscore the importance of accounting for exposure intensity when interpreting VE, they also highlight the need for a cautious interpretation that acknowledges the multifactorial nature of these relationships.

This study has several limitations. First, we categorized VE against infection from both surveillance-based test-negative design (TND) studies and database-based studies as VE against infection. These two study designs are not entirely comparable, which may affect interpretation. Second, the TND studies reviewed were observational in nature. Although many studies adjusted for confounders such as age, sex, healthcare worker status, and pre-existing conditions, residual confounding cannot be ruled out. While we conducted bias assessments to evaluate whether confounding, measurement errors, or selection bias were adequately addressed, unidentified biases may still be present. Lastly, we used the values of three indices (SI, CHI, and GRI) in the study countries as proxies for the intensity of PHSMs and exposure levels. These indices are unlikely to capture the full complexity of PHSMs, resulting in measurement errors and subsequent residual confounding.

In conclusion, our study suggests that exposure intensity to SARS-CoV-2, as reflected by PHSMs (SI, CHI, and GRI), substantially influence observed VE estimates. Stricter PHSMs correlated with higher VE estimates, while VE tended to be lower in settings with intense exposure. To provide more reliable VE estimates, future studies should account for exposure intensity and adjust for the impact of PHSMs, particularly as the virus continues to evolve and new variants emerge. For policy makers, this implies that VE estimates from areas with stringent PHSMs may overstate the vaccine’s true protective effect compared to regions with higher exposure intensity. Therefore, it is crucial to adjust VE estimates for local exposure conditions when developing vaccine strategies.

## METHODS

### Search strategy and selection criteria

This systematic review adhered to the guidelines set forth by the Preferred Reporting Items for Systematic Reviews and Meta-Analyses (PRISMA) statement (107). We conducted a standardized search in PubMed, Embase and Web of Science, using the search term “(“test negative” OR “effectiveness”) AND (“vaccine”) AND (“COVID-19” OR “SARS-CoV-2”)”. Duplicates identified across the databases were removed. The search was performed on 04 Sept 2023, without language restrictions. Additionally, we examined the reference lists of identified articles to locate further relevant studies. Two independent reviewers (XH and ZC) completed title and full-text screening and extracted data from the included studies. Any disagreements were resolved through consensus with a third reviewer (TKT).

The inclusion criteria focused on studies employing a test-negative design (TND) where all cases and controls were tested (108, 109), and where vaccine effectiveness (VE) was estimated for at least two distinct time periods to assess potential waning effects. We included published TND studies which drew participants from the general population that received a complete primary vaccination series (two doses for most vaccines; one dose for Janssen). We considered the following outcomes: (1) positive test result, (2) symptomatic disease, (3) hospitalization, (4) ICU admission, (5) severe COVID-19, or (6) death. Studies were excluded if they met any of the following criteria: (1) participants were recruited from specific sub-populations, such as healthcare professionals; (2) VE estimates were reported for only single time period; (3) studies that merely summarized or reanalysed previously-published data; (4) studies that reported only pooled VE estimates across different vaccines; (5) the study was a preprint, as these are not peer-reviewed; or (6) the full text was not available.

Data were extracted from the included studies using a standardized data collection form (Table S9), which gathered information on the following aspects: (1) study period; (2) geographic region(s); (3) study population; (4) use of clinical criteria for participant enrolment; (5) inclusion of individuals with prior SARS-CoV-2 infection; and (6) time intervals used to assess vaccine effectiveness (VE) after vaccination. For each study, we extracted confounder-adjusted VE estimates with confidence intervals separately for each endpoint (e.g., infection, hospitalization), as well as for specific vaccines and circulating virus variants. VE estimates were collected for the earliest available time interval, starting at least 14 days post-vaccination, given that antibody levels have been shown to peak by that time in previously unexposed individuals (110). In cases where studies reported multiple estimates (e.g., by age group or vaccine type), all subgroup-specific estimates were included, while overall estimates were excluded.

The quality of the studies was assessed using the Risk of Bias in Non-Randomized Studies of Interventions (ROBINS-I) tool (Sterne et al., 2016). Additionally, the certainty of the evidence for studies included in the meta-analysis was graded using the Grading of Recommendations Assessment, Development, and Evaluation (GRADE) approach (**111**). Sensitivity analyses were conducted by repeating all meta-analyses and meta-regressions while excluding studies classified as having a serious or critical risk of bias.

Our previous reviews (8, 9) indicated that studies excluding individuals with prior infections or not using clinical definitions may report artificially high VE estimates. Therefore, additional sensitivity analyses were performed by repeating all meta-analyses and meta-regressions while excluding such studies.

### Exposure intensity

The primary focus of the study was to examine the relationship between VE and potential exposure intensity. For a leaky vaccine, we hypothesized that more frequent or more intense exposures would be associated with reduced VE (Figure S1). Exposure intensity was approximated using indices from the Oxford COVID-19 Government Response Tracker (OxCGRT, https://github.com/OxCGRT/covid-policy-tracker) (112). This tracker compiled publicly accessible data on various government responses to COVID-19 and aggregated them into systematic indices. Three specific indices were extracted: (1) the Stringency Index (SI), which captures changes in school and workplace closures, containment measures, and public information campaigns; (2) the Containment and Health Index (CHI), which includes SI data along with changes in health policy; and (3) the Government Response Index (GRI), which provides a comprehensive measure encompassing SI, CHI, and economic support measures to mitigate the pandemic’s impact on economic activities.

The scale of each index was 0 to 100, with higher scores reflecting stricter government policies, and thus, shorter durations of high exposure intensity. We used the average of these indices during the study period to measure the average control intensity and approximate the likely exposure intensity experienced by participants in each study (Figure S2).

### Meta-analysis

In all the included studies, VE was defined as 100%*(1-OR). VE estimates were transformed to the odds ratio (OR) scale, analyzed via meta-analysis, and back-transformed to the VE scale for interpretation. The pooled OR was calculated using random-effects meta-analyses, employing the inverse variance method along with the restricted maximum likelihood estimator to account for heterogeneity (113-116). Heterogeneity was evaluated using Cochran’s **Q** and the **I**^2^ statistic (117). An **I**^2^ value exceeding 75% was interpreted as indicative of high heterogeneity (118). Additionally, sensitivity analysis was performed using fixed-effects meta-analysis.

Severe disease was defined as hospitalization, ICU admission, or death. VE estimates not limited to severe cases were classified as VE against infection, referring to VE against positive test results or symptomatic infection without hospitalization.

Pooled VE estimates were stratified by the tertile of each of the three indices, to explore their relationship with VE estimates. Pooled VE estimates were further stratified by the predominant circulating virus and the type of vaccine administered. Most studies did not provide variant-specific VE estimates but instead reported study periods and the general prevalence of variants during those times. Thus, VE estimates were grouped based on the predominant circulating virus as follows: (1) Omicron BA.4/BA.5 or later, (2) Omicron BA.1/BA.2, (3) late-Delta (a period with Delta and Omicron co-circulation), and (4) Delta and pre-Delta, which included ancestral strains and earlier variants. Vaccine types were categorized into: (1) mRNA vaccines (Moderna, Pfizer-BioNTech), (2) adenovirus vector vaccines (AstraZeneca, Janssen, Gamaleya), and (3) inactivated virus vaccines (Sinovac Biotech, Sinopharm).

### Meta-regression

We used meta-regression to assess the impact of exposure intensity, approximated by the level of public health and social measures as indicated by the Government Response Index (GRI), on vaccine effectiveness (VE) estimates. Initially, correlation analyses were performed using Pearson (r) and Spearman (*ρ*) correlation coefficients, to determine the association between these indices and VE estimates. The meta-regression models were adjusted for several covariates: age group (age <65 or ≥65 years), vaccine types, predominant circulating virus strain, inclusion/exclusion of participants with prior infections, and the use of clinical criteria for participant enrolment.

The fitted meta-regression model estimated the ratio of odds ratios (ROR) for each 10-unit increase in the indices. On the OR scale, values closer to 0 indicated greater vaccine effectiveness, while values closer to 1 suggested reduced effectiveness. This interpretation contrasts with the VE scale, where values closer to 0 imply lower effectiveness. For instance, if the ROR for the GRI is less than 1, it suggests that studies with a higher average GRI during their study period reported lower ORs (indicating greater vaccine effectiveness) compared to studies with lower average GRI scores. On the VE scale, this corresponds to higher VE estimates for studies conducted in regions with more stringent government responses during the study period.

All statistical analyses were performed using R version 4.0.5 (R Foundation for Statistical Computing, Vienna, Austria), employing the metafor package for meta-analyses and the robvis package for visualizing risk of bias assessments.

## Data Availability

All data produced in the present study are available upon reasonable request to the authors

## Author Contributions

TKT, SGS and BJC contributed to the study conception and design. Material preparation, data collection and analysis were performed by TKT, XH, CW and YW. The first draft of the manuscript was written by TKT and all authors commented on previous versions of the manuscript. All authors read and approved the final manuscript.

## Ethics approval

This is a systematic review and meta-analysis. No ethical approval is required.

