## Appendix for "Intensity of public health and social measures are associated with effectiveness of SARS-CoV-2 vaccine in test-negative study"

### 1 Supplementary Note 1. Further Information on meta-analyses.

Our meta-analysis revealed that pooled VE against infection varied by vaccine type: 63% (95% CI: 61%, 66%) for mRNA vaccines, 66% (95% CI: 61%, 71%) for adenovirus vector vaccines, and 40% (95% CI: 34%, 45%) for inactivated virus vaccines (Figure S3). When stratified by circulating virus, VE against infection was markedly lower during the Omicron BA.4/5 period and beyond (19%; 95% CI: 14%, 24%) and during the Omicron BA.1/2 period (28%; 95% CI: 25%, 31%), compared to the late-Delta period (80%; 95% CI: 78%, 81%) and the pre-Delta/Delta period (81%; 95% CI: 79%, 83%). Similarly, VE against severe disease declined during the Omicron BA.4/5 period (33%; 95% CI: 27%, 38%) and the Omicron BA.1/2 period (61%; 95% CI: 57%, 65%) relative to the pre-Delta/Delta period (91%; 95% CI: 89%, 92%) and the late-Delta period (93%; 95% CI: 92%, 94%). These trends were consistent when using fixed-effects models (Figure S7).

Studies including participants with prior COVID-19 infection reported higher pooled VE estimates against infection (68%; 95% CI: 66%, 70%) compared to those excluding such participants (55%; 95% CI: 51%, 59%). Similarly, pooled VE against severe disease was higher in studies including participants with prior infection (88%; 95% CI: 86%, 89%) than in those excluding them (84%; 95% CI: 81%, 87%), with significant heterogeneity across estimates (I²=100%). Further stratification by enrollment criteria indicated that pooled VE against infection was higher in studies not requiring clinical case definitions for inclusion (71%; 95% CI: 69%, 74%) compared to those applying clinical case definitions (57%; 95% CI: 54%, 59%). This pattern was also observed for VE against severe disease, which was higher in studies without clinical case definitions (92%; 95% CI: 90%, 93%) than in those with such criteria (81%; 95% CI: 79%, 83%), again with considerable heterogeneity (I²=100%).

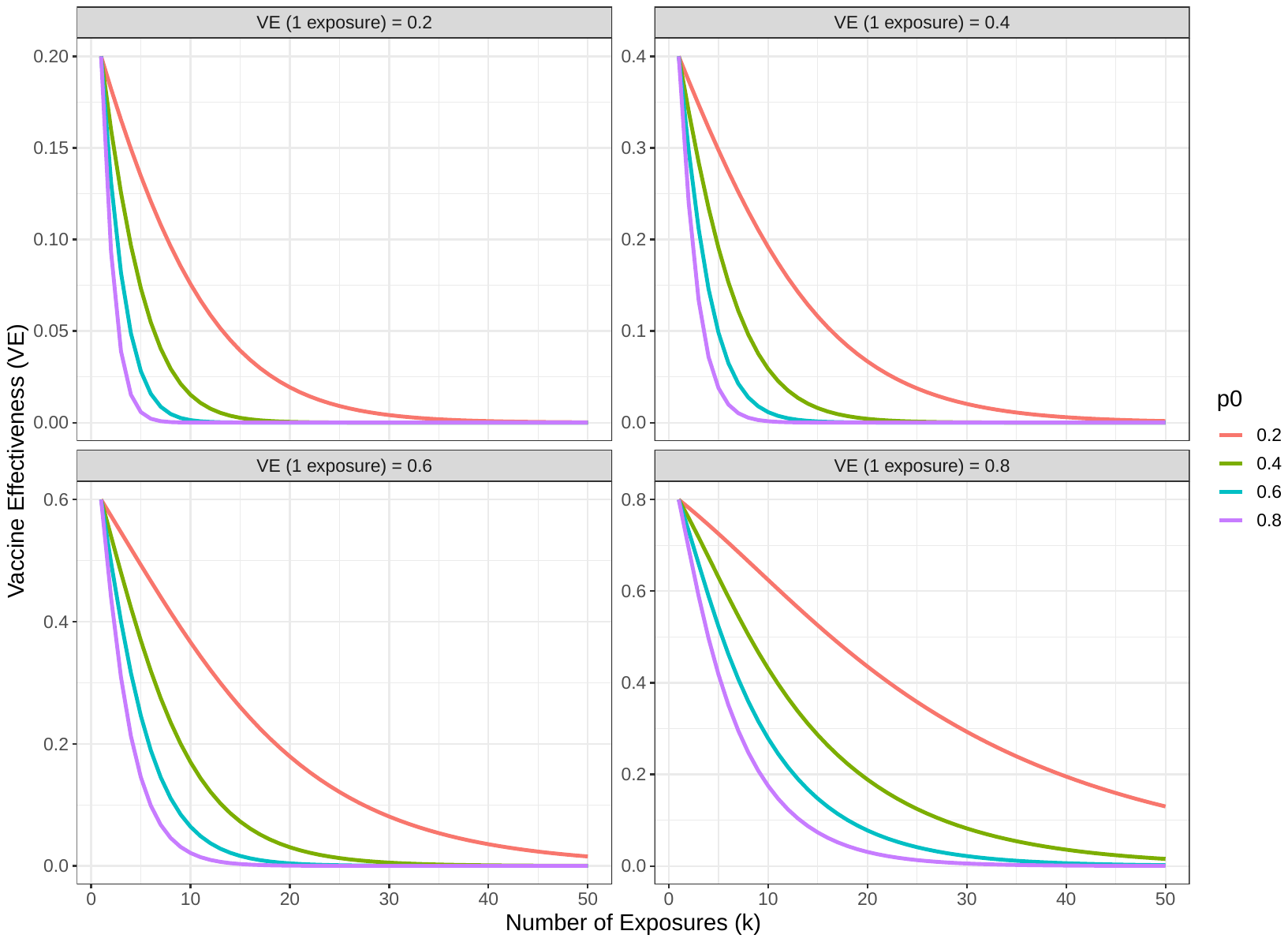

### Figure S1. Vaccine Effectiveness (VE) as a Function of Number of Exposures (k) under Different Initial VE Values.

The figure illustrates the relationship between the number of exposures (k) and vaccine effectiveness (VE) for varying baseline VE levels after a single exposure (VE(1)). Panels represent different scenarios of VE(1), with values of 0.2, 0.4, 0.6, and 0.8. Vaccine effectiveness diminishes with an increasing number of exposures, with the rate of decline varying based on initial effectiveness. p0p0 represents the proportion susceptible after one exposure. Axes are labeled with the number of exposures (kk) on the x-axis and vaccine effectiveness (VE) on the y-axis, highlighting the interplay between exposure frequency and vaccine performance

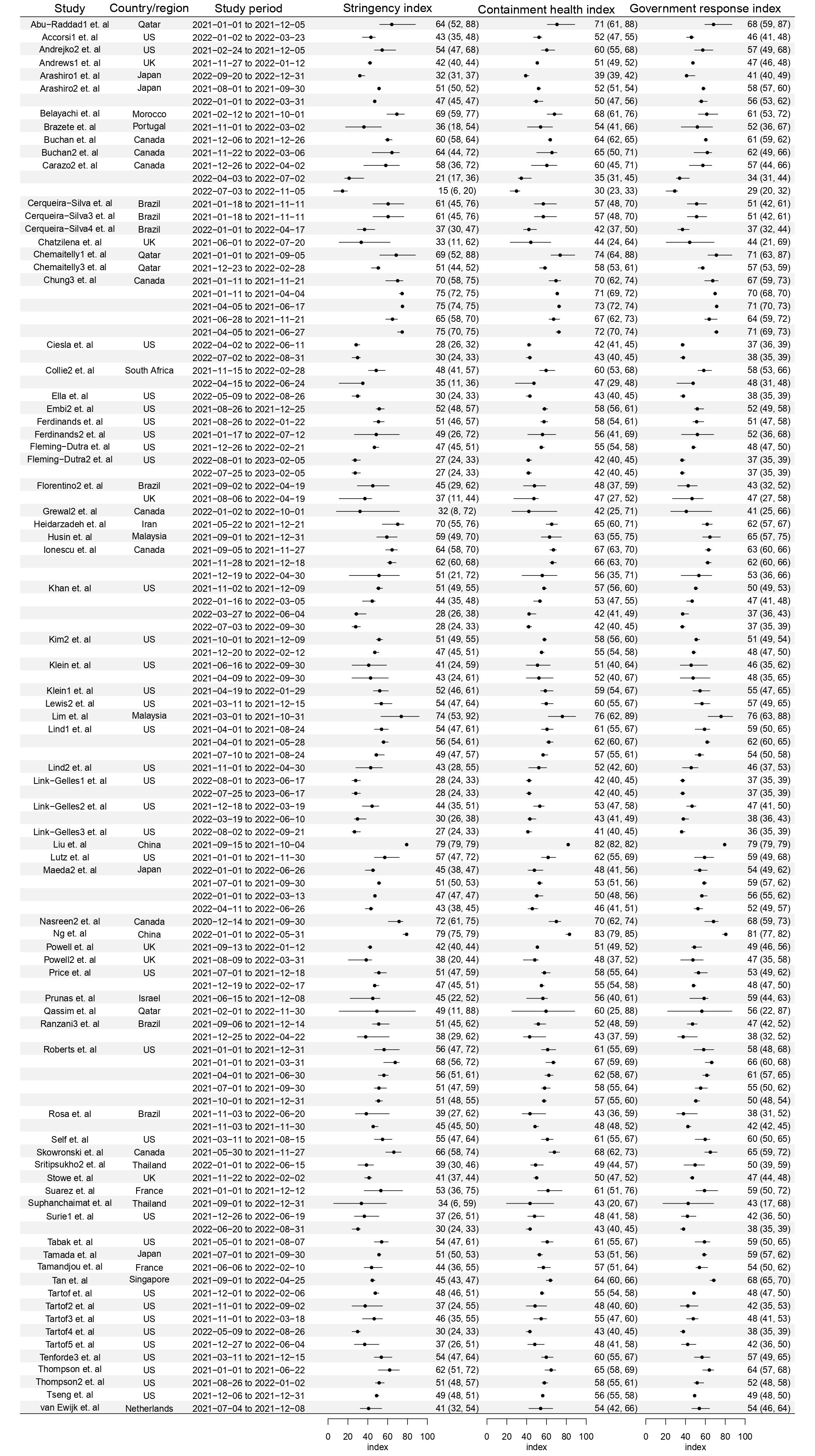

Figure S2: Average of government response index during study for each identified study

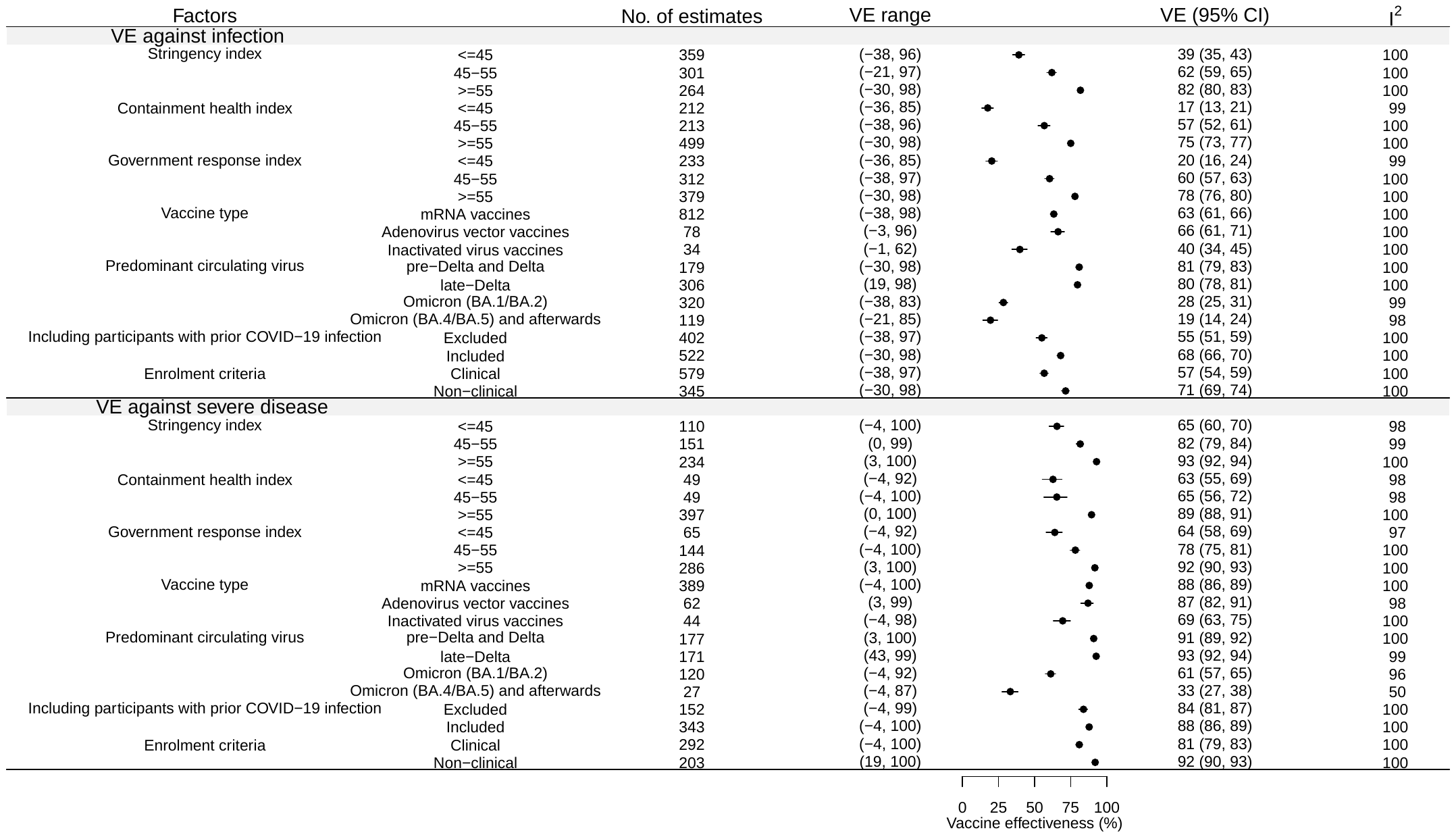

Figure S3. Pooled vaccine effectiveness (VE) estimates by key factors. Meta-analysis results displaying pooled VE estimates against infection and severe disease, stratified by vaccine type, predominant circulating variant, inclusion of prior infection, and enrolment criteria. Substantial heterogeneity in VE is observed across subgroups, with VE estimates higher in settings with stricter control measures (higher index values) and among participants with prior infection.

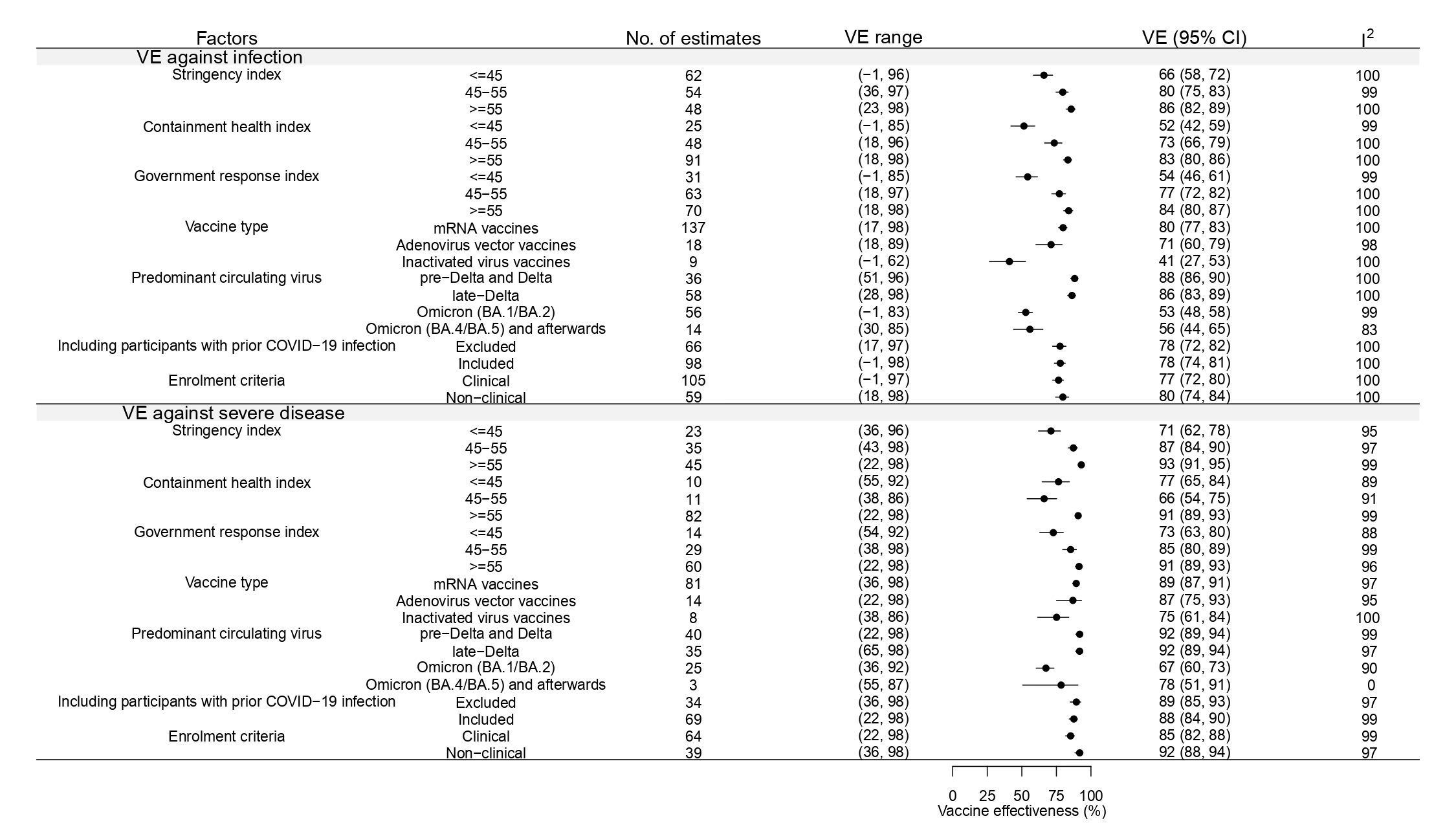

Figure S4. Pooled VE estimates reported for the first interval after vaccination against infection and severe disease by each index measuring the intensity of control measures, circulating virus, vaccine types, the inclusion or exclusion of participants with prior COVID-19 infection and enrolment criteria from random-effect meta-analysis.

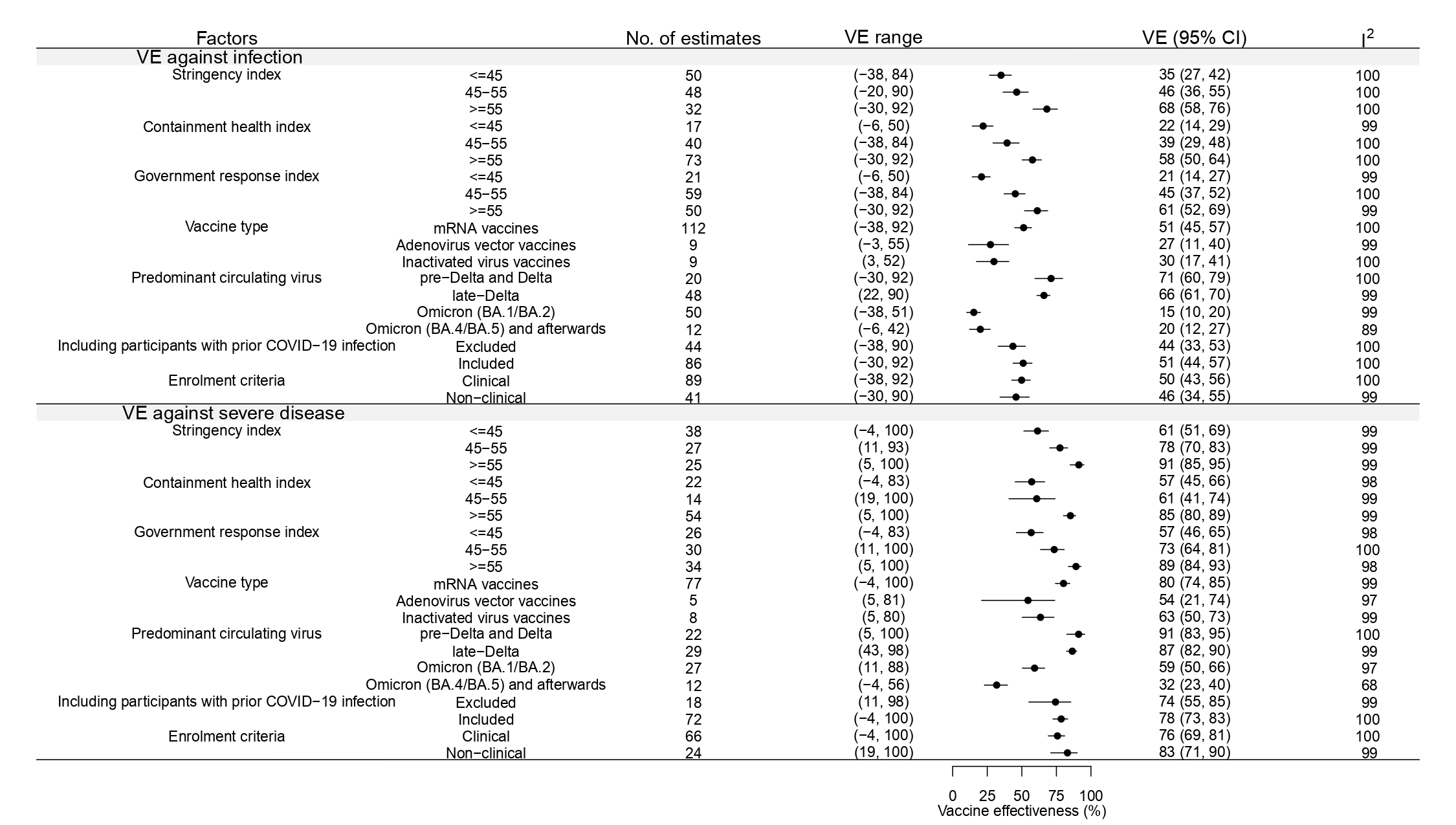

Figure S5. Pooled VE estimates reported for the last interval after vaccination against infection and severe disease by each index, circulating virus, vaccine types, the inclusion or exclusion of participants with prior COVID-19 infection and enrolment criteria from random-effect meta-analysis.

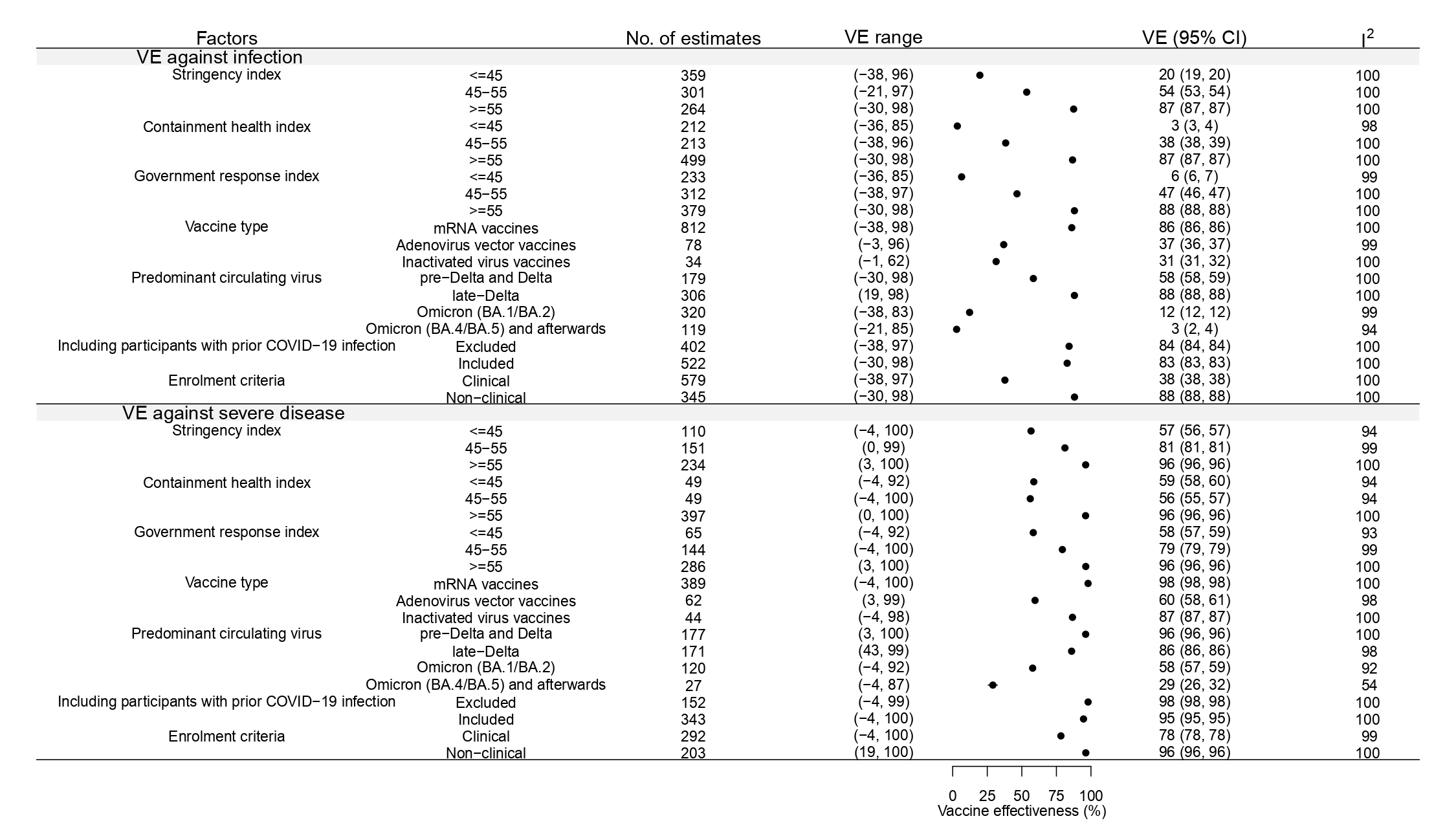

Figure S6. Pooled VE estimates reported for the last interval after vaccination against infection and severe disease by each index, circulating virus, vaccine types, the inclusion or exclusion of participants with prior COVID-19 infection and enrolment criteria from fixed-effect meta-analysis.

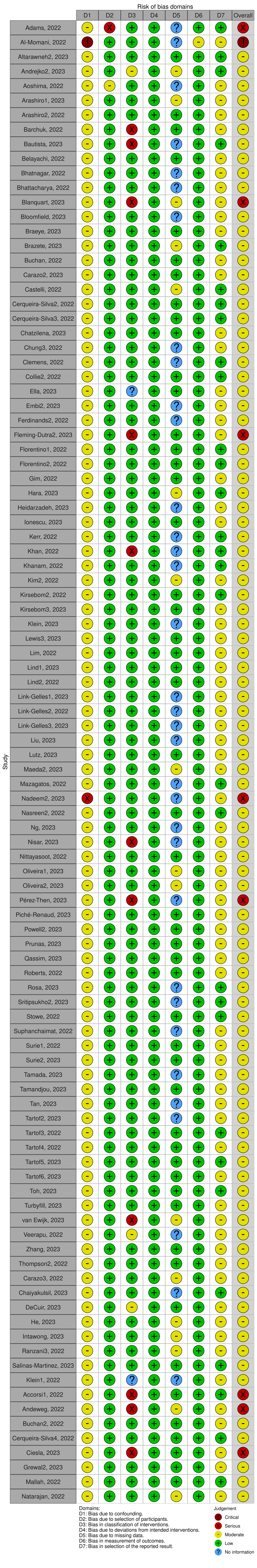

Figure S7. Bias assessment using the Risk Of Bias In Non-randomized Studies – of Interventions (ROBINS-I).

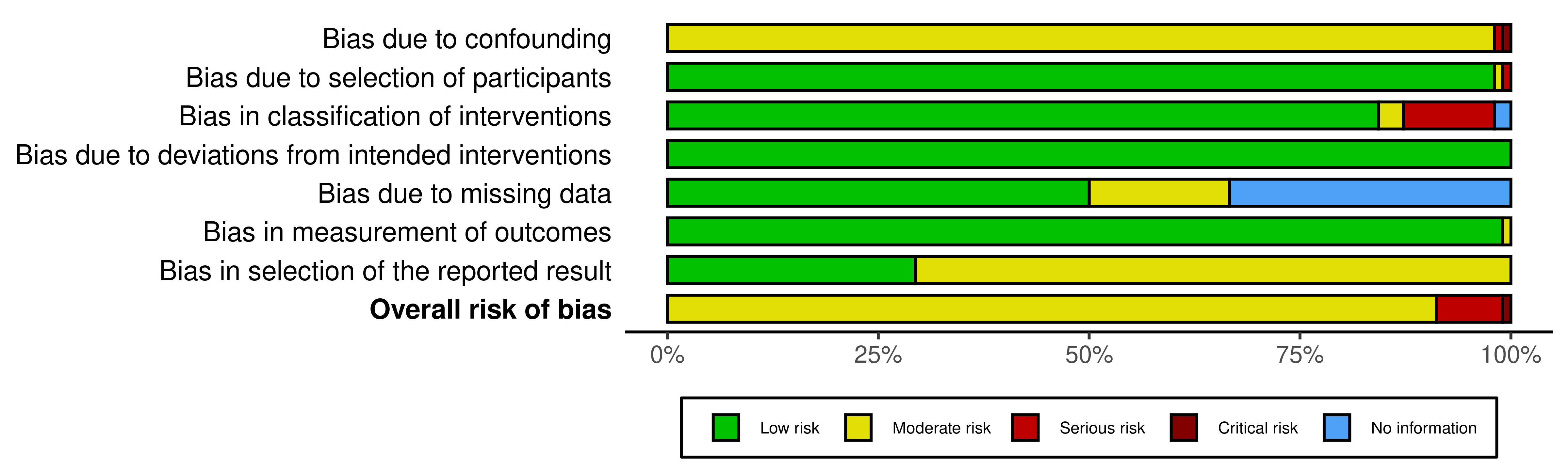

Figure S8. Summary of bias assessment using the Risk Of Bias In Non-randomized Studies – of Interventions (ROBINS-I).

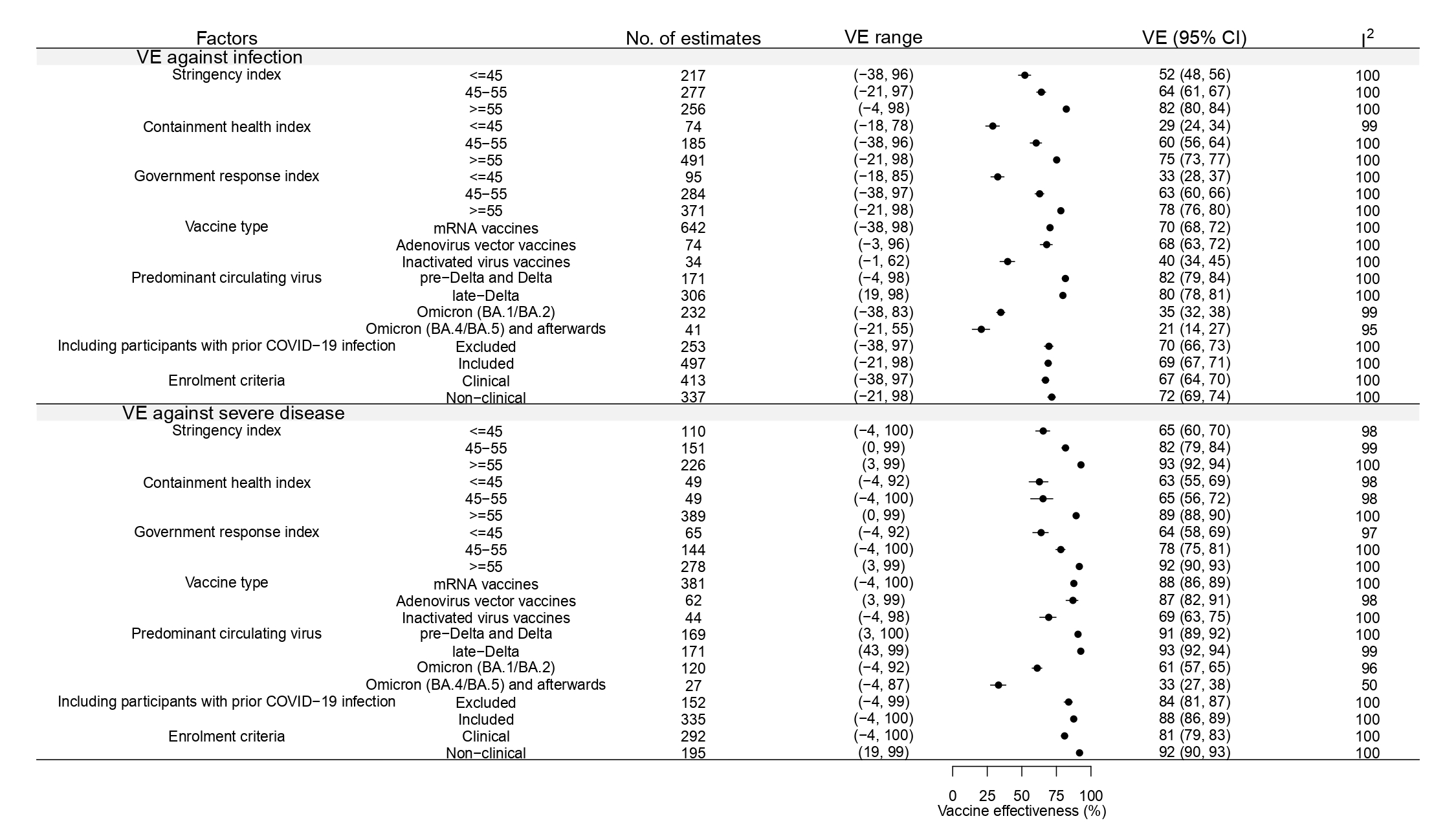

Figure S9. Pooled VE estimates reported for the last interval after vaccination against infection and severe disease by each index, circulating virus, vaccine types, the inclusion or exclusion of participants with prior COVID-19 infection and enrolment criteria, excluding estimates from studies classified as having serious or critical bias.

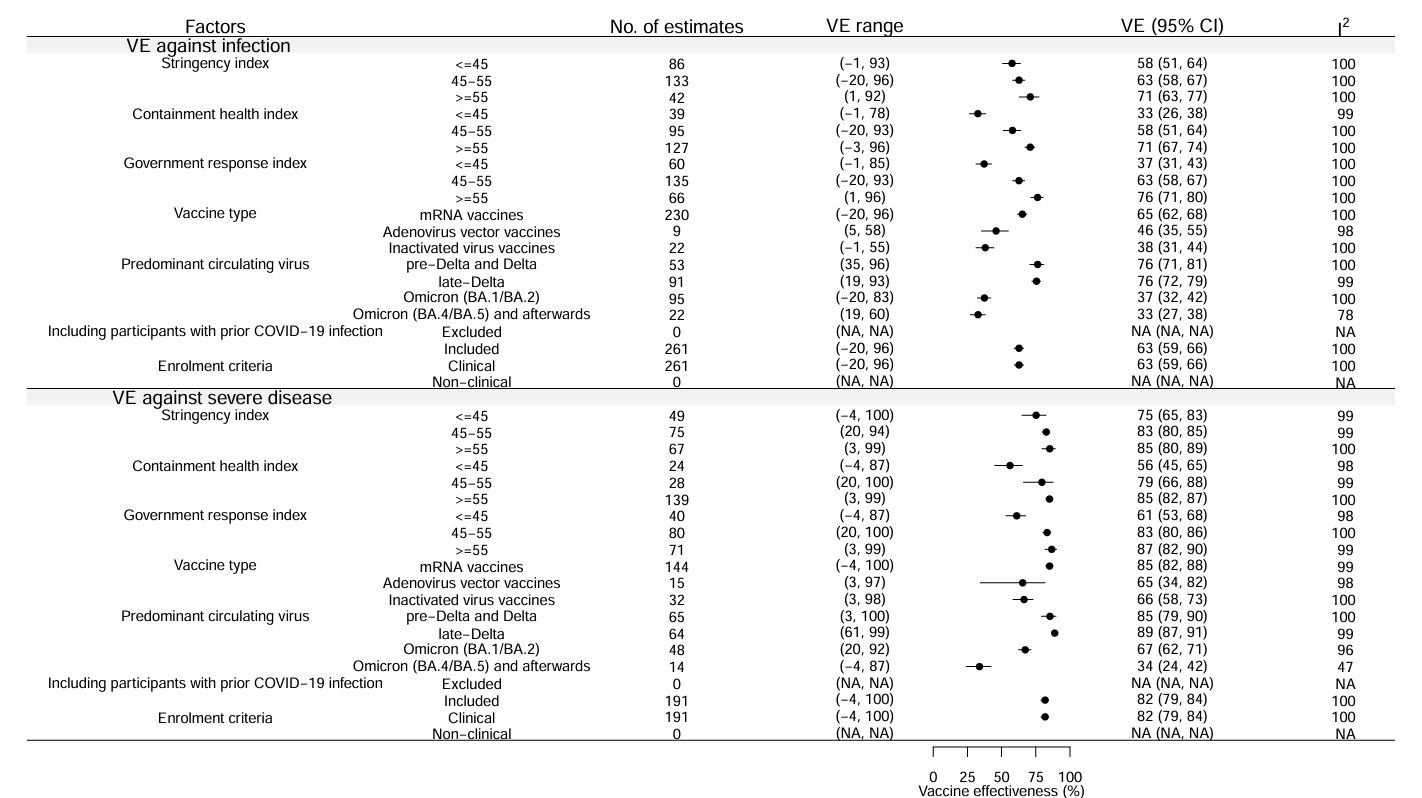

Figure S10. Pooled VE estimates reported for the last interval after vaccination against infection and severe disease by each index, circulating virus, vaccine types, excluding from studies not using clinical case definition or excluding participants with prior infections.

Table S1. Pearson and spearman correlation for each containment index by endpoint and predominant circulating variants.

|  |  | stringency index | | containment health index | | government response index | |
| --- | --- | --- | --- | --- | --- | --- | --- |
|  |  | pearson correlation | spearman correlation | pearson correlation | spearman correlation | pearson correlation | spearman correlation |
| infection | Overall (black) | 0.62 (0.57, 0.66) | 0.62 (0.57, 0.66) | 0.57 (0.52, 0.62) | 0.60 (0.55, 0.64) | 0.57 (0.52, 0.62) | 0.57 (0.53, 0.62) |
|  | pre-delta and delta (blue) | 0.00 (-0.11, 0.13) | 0.04 (-0.09, 0.17) | 0.02 (-0.15, 0.23) | 0.10 (-0.07, 0.27) | 0.10 (-0.08, 0.30) | 0.14 (-0.02, 0.30) |
|  | late-delta (purple) | 0.36 (0.25, 0.46) | 0.30 (0.18, 0.41) | 0.32 (0.19, 0.44) | 0.26 (0.14, 0.36) | 0.28 (0.13, 0.41) | 0.22 (0.09, 0.34) |
|  | Omicron (BA.1/BA.2) (gold) | 0.33 (0.23, 0.43) | 0.30 (0.19, 0.41) | 0.22 (0.12, 0.32) | 0.22 (0.11, 0.34) | 0.27 (0.17, 0.36) | 0.28 (0.16, 0.40) |
|  | Omicron (BA.4/BA.5) and afterwards (green) | -0.26 (-0.36, -0.16) | -0.25 (-0.42, -0.09) | -0.27 (-0.37, -0.18) | -0.33 (-0.49, -0.18) | -0.23 (-0.33, -0.11) | -0.24 (-0.41, -0.08) |
| severe disease | Overall (black) | 0.43 (0.33, 0.52) | 0.50 (0.41, 0.58) | 0.45 (0.37, 0.53) | 0.57 (0.50, 0.64) | 0.43 (0.35, 0.51) | 0.51 (0.44, 0.58) |
|  | pre-delta and delta (blue) | -0.21 (-0.31, -0.10) | 0.03 (-0.15, 0.19) | 0.08 (-0.01, 0.17) | 0.30 (0.14, 0.43) | 0.15 (0.06, 0.23) | 0.36 (0.21, 0.49) |
|  | late-delta (purple) | 0.42 (0.30, 0.54) | 0.52 (0.40, 0.63) | 0.40 (0.25, 0.53) | 0.53 (0.41, 0.64) | 0.37 (0.22, 0.50) | 0.45 (0.31, 0.57) |
|  | Omicron (BA.1/BA.2) (gold) | 0.15 (-0.02, 0.29) | 0.16 (-0.03, 0.31) | -0.01 (-0.17, 0.13) | 0.02 (-0.18, 0.19) | -0.06 (-0.21, 0.09) | -0.01 (-0.21, 0.15) |
|  | Omicron (BA.4/BA.5) and afterwards (green) | 0.14 (-0.20, 0.44) | 0.26 (-0.12, 0.57) | 0.08 (-0.30, 0.38) | 0.00 (-0.37, 0.37) | 0.18 (-0.16, 0.48) | 0.26 (-0.12, 0.57) |

Table S2. Summary of included studies in the systematic review and meta-analysis.

| **Author (year)** | **Participant** | **Study period** | **Location** | **Methods to determine vaccination status** | **Periods of diagnosis tests** | **Types of diagnosis tests** | **Timing since vaccination included** | **Method of determining prior infection** | **Vaccine type** | **Circulating Virus type** | **Endpoint (grouped endpoint in our analysis)** | **Recruitment criteria** |
| --- | --- | --- | --- | --- | --- | --- | --- | --- | --- | --- | --- | --- |
| Abu-Raddad1 (2022) (1) | general | 2021-01-01 to 2021-12-05 | Qatar | Documented | As of 2021/12/05 | PCR-positive swab, regardless of the presence of symptoms | 0-30 & 61-90 & 91-120 & 121-180 & 181+ | Not available (NA) | mRNA | Pre-Delta and Delta | Infection (infection) & hospitalization and death (severe disease) | Non-clinical |
| Accorsi1 (2022) (2) | >=18 years of age | 2022-01-02 to 2022-03-23 | US | Self-reported | From January 2, 2022, to March 23, 2022 | NAAT | 31-60 & 91-120 | Documented | Adenovirus vector vaccines | Omicron (BA.1/BA.2) | Symptomatic (infection) | Clinical |
| Andrejko2 (2023) (3) | >=13 years of age | 2021-02-24 to 2021-12-05 | US | Self-reported | Same as study period,between February 24 and December 5, 2021 | molecular, antigen, or serological | 0-30 & 31-60 & 91-120 & 121-180 & 181+ | Molecular, antigen, or serological | mRNA | Pre-Delta and Delta | Symptomatic (infection) | Non-clinical |
| Andrews1 (2022) (4) | >=18 years of age | 2021-11-27 to 2022-01-12 | UK | Documented | Same as study period, November 27, 2021-January 12, 2022 | PCR | 31-60 & 61-90 & 121-180 & 181+ | PCR | mRNA & Adenovirus vector vaccines | Late-Delta & Omicron (BA.1/BA.2) | Symptomatic (infection) | Clinical |
| Arashiro1 (2023) (5) | >=16 years of age | 2022-09-20 to 2022-12-31 | Japan | Self-reported plus documented | Same as study period,2022-09-20 to 2022-12-31 | PCR | 31-60 & 181+ | Self-reported | mRNA | Omicron (BA.4/BA.5) and afterwards | Symptomatic (infection) | Clinical |
| Arashiro2 (2022) (6) | >=20 years of age | 2021-08-01 to 2021-09-30 & 2022-01-01 to 2022-03-31 | Japan | Self-reported plus documented | Between 1 August 2021 and 31 March 2022 | PCR | 31-60 & 181+ | NA | mRNA | Late-Delta & Omicron (BA.1/BA.2) | Symptomatic (infection) | Clinical |
| Belayachi (2022) (7) | >=18 years | 2021-02-12 to 2021-10-01 | Morocco | Documented | Between February 1 2021 and October 1 2021 | RT-PCR | 0-30 & 61-90 & 91-120 & 121-180 & 181+ | NA | Inactivated virus vaccines | Pre-Delta and Delta | Hospitalization (severe disease) | Non-clinical |
| Brazete (2023) (8) | >=18 years of age | 2021-11-01 to 2022-03-02 | Portugal | Documented | Same as study period, 1 November 2021 to 2 March 2022 | rRT-PCR | 91-120 & 181+ | NA | mRNA & Adenovirus vector vaccines | Omicron (BA.1/BA.2) | Symptomatic (infection) | Clinical |
| Buchan (2022) (9) | >=18 years of age | 2021-12-06 to 2021-12-26 | Canada | Documented | Between December 6 and 26,2021 | RT-PCR | 31-60 & 91-120 & 181+ | NA | mRNA | Omicron (BA.1/BA.2) & late-Delta | Symptomatic (infection) & severe (severe disease) | Clinical |
| Buchan2 (2022) (10) | 12-17 years of age | 2021-11-22 to 2022-03-06 | Canada | Documented | Between November 22, 2021 and March 6, 2022 | RT-PCR | 31-60 & 91-120 & 181+ | NA | mRNA | Omicron (BA.1/BA.2) & late-Delta | Symptomatic (infection) & severe (severe disease) | Non-clinical |
| Carazo2 (2023) (11) | >=60 years of age | 2021-12-26 to 2022-04-02 & 2022-04-03 to 2022-07-02 & 2022-07-03 to 2022-11-05 | Canada | Documented | Between Dec 26, 2021, and Nov 5, 2022 | NAAT | 31-60 & 61-90 & 121-180 & 181+ | NAAT | mRNA | Omicron (BA.1/BA.2) & Omicron (BA.4/BA.5) and afterwards | Hospitalization (severe disease) | Clinical |
| Cerqueira-Silva (2022) (12) | >=18 years of age | 2021-01-18 to 2021-11-11 | Brazil | Documented | Between 18 January 2021 and 11 November 2021 | RT–PCR and antigen | 0-30 & 61-90 & 91-120 & 121-180 & 181+ | NA | Inactivated virus vaccines | Pre-Delta and Delta | Symptomatic (infection) & Hospitalization or death (severe disease) | Clinical |
| Cerqueira-Silva3 (2022) (13) | >=18 years of age | 2021-01-18 to 2021-11-11 | Brazil | Documented | From 24 February 2020 to 11 November 2021 | RT-PCR | 0-30 & 61-90 & 91-120 & 121-180 & 181+ | Test | Inactivated virus vaccines | Pre-Delta and Delta | Infection (infection) & hospitalization or death (severe disease) | Clinical |
| Cerqueira-Silva4 (2022) (14) | >=18 years of age | 2022-01-01 to 2022-04-17 | Brazil | Documented | Between January 01, 2022, and April 17, 2022 | RT-PCR / Lateral-flow test | 91-120 & 181+ | NA | Inactivated virus vaccines | Omicron (BA.1/BA.2) | Symptomatic (infection) & severe (severe disease) | Clinical |
| Chatzilena (2023) (15) | >=18 years of age | 2021-06-01 to 2022-07-20 | UK | Documented | Same as study period, 2021-06-01 to 2022-07-20 | PCR | 31-60 & 181+ | Symptoms | mRNA | Late-Delta | Hospitalization or death (severe disease) | Clinical |
| Chemaitelly1 (2021) (16) | >=12 years of age | 2021-01-01 to 2021-09-05 | Qatar | Documented | January 1, 2021 to September 5, 2021 | PCR | 31-60 & 91-120 & 121-180 & 181+ | NA | mRNA | Pre-Delta and Delta | Infection (infection) & hospitalization and death (severe disease) | Non-clinical |
| Chemaitelly3 (2022) (17) | general | 2021-12-23 to 2022-02-28 | Qatar | Documented | December 23, 2021, December 23, 2021 and February 28, 2022 | PCR | 61-90 & 181+ | Documented (sensitivity analysis) | mRNA | Omicron (BA.1/BA.2) | Symptomatic (infection) | Non-clinical |
| Chung3 (2022) (18) | >=16 years of age | 2021-01-11 to 2021-11-21 & 2021-01-11 to 2021-04-04 & 2021-04-05 to 2021-06-17 & 2021-06-28 to 2021-11-21 & 2021-04-05 to 2021-06-27 | Canada | Documented | Between January 11, 2021 and November 21, 2021 | RT-PCR | 31-60 & 91-120 & 181+ | Documented | mRNA & Adenovirus vector vaccines | Pre-Delta and Delta & late-Delta | Any infection (infection) & severe (severe disease) | Non-clinical |
| Ciesla (2023) (19) | >=5 years of age | 2022-04-02 to 2022-06-11 & 2022-07-02 to 2022-08-31 | US | Self-reported | April 2 to June 11, 2022 and July 2 to August 31, 2022 | Nucleic acid amplification tests (NAATs) | 0-30 & 31-60 & 61-90 & 91-120 & 121-180 & 181+ | Testing | mRNA | Omicron (BA.1/BA.2) & Omicron (BA.4/BA.5) and afterwards | Symptomatic (infection) | Clinical |
| Collie2 (2022) (20) | >=18 years of age | 2021-11-15 to 2022-02-28 & 2022-04-15 to 2022-06-24 | South Africa | Documented | Same as study period, from November 15, 2021 to June 24, 2022 | PCR | 0-30 & 61-90 & 121-180 & 181+ | NA | mRNA | Omicron (BA.1/BA.2) & Omicron (BA.4/BA.5) and afterwards | Hospitalization (severe disease) | Non-clinical |
| Ella (2023) (21) | >=18 years of age | 2022-05-09 to 2022-08-26 | US | NA | Between May 9 and Aug 26, 2022 | PCR | 61-90 & 91-120 & 181+ | NA | mRNA | Omicron (BA.4/BA.5) and afterwards | Outpatient (infection) & hospitalization (severe disease) | Clinical |
| Embi2 (2023) (22) | >=18 years of age | 2021-08-26 to 2021-12-25 | US | Documented | From 26 August-25 December 2021 | Molecular assays | 61-90 & 121-180 | Testing | mRNA | Late-Delta | Emergency department and urgent care encounters (infection) & hospitalization (severe disease) | Clinical |
| Ferdinands (2022) (23) | >=18 years of age | 2021-08-26 to 2022-01-22 | US | Documented | Same as study period, August 2021–January 2022 | PCR | 31-60 & 61-90 & 91-120 & 121-180 & 181+ | NA | mRNA | Late-Delta & Omicron (BA.1/BA.2) | Emergency department and urgent care encounters (infection) & hospitalization (severe disease) | Clinical |
| Ferdinands2 (2022) (24) | >=18 years of age | 2021-01-17 to 2022-07-12 | US | Documented | 17 January to 3 May 2021 | Molecular testing | 31-60 & 91-120 & 181+ | NA | mRNA | Pre-Delta and Delta & late-Delta & Omicron (BA.1/BA.2) | Emergency department and urgent care encounters (infection) & hospitalization (severe disease) | Clinical |
| Fleming-Dutra (2022) (25) | 5-15 years of age | 2021-12-26 to 2022-02-21 | US | Self-reported | From December 26, 2021, to February 21, 2022 | NAATs | 0-30 & 31-60 & 61-90 & 91-120 & 121-180 & 181+ | Positive test result | mRNA | Omicron (BA.1/BA.2) | Symptomatic (infection) | Clinical |
| Fleming-Dutra2 (2023) (26) | 3-5 years of age | 2022-08-01 to 2023-02-05 & 2022-07-25 to 2023-02-05 | US | Self-reported | During September 19, 2022–February 5, 2023 | NAATs | 31-60 & 121-180 | NA | mRNA | Omicron (BA.4/BA.5) and afterwards | Symptomatic (infection) | Clinical |
| Florentino2 (2022) (27) | 12-17 years of age | 2021-09-02 to 2022-04-19 & 2021-08-06 to 2022-04-19 | Brazil & UK | Documented | From Sept 2, 2021, to April 19, 2022 | RT-PCR or antigen test | 0-30 & 31-60 & 61-90 & 91-120 & 121-180 & 181+ | Previous confirmed infection | mRNA | Late-Delta & Omicron (BA.1/BA.2) | Symptomatic (infection) & severe (severe disease) | Clinical |
| Grewal2 (2023) (28) | >=50 years of age | 2022-01-02 to 2022-10-01 | Canada | Documented | From January 2 to October 1, 2022 | RT-PCR | 181+ | RT-PCR | mRNA | Omicron (BA.1/BA.2) & Omicron (BA.4/BA.5) and afterwards | Severe (severe disease) | Clinical |
| Heidarzadeh (2023) (29) | >=5 years of age | 2021-05-22 to 2021-12-21 | Iran | Documented | Same as study period, from May 22 to December 21, 2021 | RT-PCR | 0-30 & 61-90 & 91-120 & 121-180 & 181+ | PCR | Adenovirus vector vaccines & Inactivated virus vaccines | Pre-Delta and Delta | Hospitalization (severe disease) | Clinical |
| Husin (2022) (30) | 12-17 years of age | 2021-09-01 to 2021-12-31 | Malaysia | Documented | September 1, 2021, until December 31, 2021 | RT-PCR or rapid antigen | 31-60 & 61-90 & 91-120 & 121-180 | NA | mRNA | Late-Delta | Infection (infection) | Non-clinical |
| Ionescu (2023) (31) | 12-17 years of age | 2021-09-05 to 2021-11-27 & 2021-11-28 to 2021-12-18 & 2021-12-19 to 2022-04-30 | Canada | Documented | Same as study period, between 5 September 2021 and 30 April 2022 | NAAT | 0-30 & 31-60 & 61-90 & 91-120 & 121-180 & 181+ | NA | mRNA | Late-Delta & Omicron (BA.1/BA.2) | Infection (infection) | Non-clinical |
| Khan (2022) (32) | 5-11 years of age | 2021-11-02 to 2021-12-09 & 2022-01-16 to 2022-03-05 & 2022-03-27 to 2022-06-04 & 2022-07-03 to 2022-09-30 | US | Self-reported | Between November 2, 2021, and September 30, 2022 | PCR | 0-30 & 31-60 & 91-120 & 121-180 & 181+ | Self-reported | mRNA | Late-Delta & Omicron (BA.1/BA.2) & Omicron (BA.4/BA.5) and afterwards | Infection (infection) | Non-clinical |
| Kim2 (2022) (33) | >=18 years of age | 2021-10-01 to 2021-12-09 & 2021-12-20 to 2022-02-12 | US | Documented | Between October 2021 and February 2022 | RT-PCR | 31-60 & 61-90 & 121-180 | self-reported | mRNA | Late-Delta & Omicron (BA.1/BA.2) | Symptomatic (infection) | Clinical |
| Klein (2023) (34) | 12-17 years of age | 2021-06-16 to 2022-09-30 & 2021-04-09 to 2022-09-30 | US | Documented | Same as study period, from April 9, 2021, to September 2022 | RT-PCR | 31-60 & 121-180 & 181+ | NA | mRNA | Pre-Delta and Delta & late-Delta | Emergency department and urgent care encounters (infection) & hospitalization (severe disease) | Clinical |
| Klein1 (2022) (35) | 5-17 years of age | 2021-04-19 to 2022-01-29 | US | NA | Same as study period, during April 9, 2021 to January 29, 2022 | RT-PCR | 31-60 & 61-90 & 181+ | NA | mRNA | Omicron (BA.1/BA.2) & late-Delta | Emergency department and urgent care encounters (infection) & hospitalization (severe disease) | Clinical |
| Lewis2 (2022) (36) | >=18 years of age | 2021-03-11 to 2021-12-15 | US | Documented | During March 11–December 15, 2021 | Nucleic acid amplification test[NAAT] or antigen test | 31-60 & 121-180 & 181+ | Self-reported | Adenovirus vector vaccines | Pre-Delta and Delta | Hospitalization (severe disease) | Non-clinical |
| Lim (2022) (37) | >=18 years of age | 2021-03-01 to 2021-10-31 | Malaysia | Documented | Between 1 March 2021 and 31 October 2021 | RT-PCR | 0-30 & 31-60 & 61-90 & 91-120 & 121-180 | NA | mRNA | Late-Delta | Infection (infection) & severe or critical disease (severe disease) | Non-clinical |
| Lind1 (2023) (38) | >=16 years of age | 2021-04-01 to 2021-08-24 & 2021-04-01 to 2021-05-28 & 2021-07-10 to 2021-08-24 | US | Documented | Between 1 April and 24 August 2021 | RT-PCR | 31-60 & 121-180 | RT-PCR or rapid antigen test | mRNA | Pre-Delta and Delta & late-Delta | Infection (infection) & hospitalization (severe disease) | Non-clinical |
| Lind2 (2022) (39) | >=5 years of age | 2021-11-01 to 2022-04-30 | US | Documented | Between November 1, 2021 and April 30, 2022 | RT-PCR | 61-90 & 181+ | Documented | mRNA | Omicron (BA.1/BA.2) | Infection (infection) | Non-clinical |
| Link-Gelles1 (2023) (40) | 6 months to 5 years of age | 2021-11-01 to 2022-04-30 & 2022-07-25 to 2023-06-17 | US | Documented | During June 20, 2022–June 3, 2023 | nucleic acid amplification testing | 31-60 & 121-180 & 181+ | Medical record | mRNA | Omicron (BA.4/BA.5) and afterwards | Emergency department and urgent care encounters (infection) | Clinical |
| Link-Gelles2 (2022) (41) | >=18 years of age | 2021-12-18 to 2022-03-19 & 2022-03-19 to 2022-06-10 | US | Documented | During December 18, 2021 to June 10, 2022 | SARS-CoV-2 molecular test | 61-90 & 121-180 | Documented | mRNA | Omicron (BA.1/BA.2) | Emergency department and urgent care encounters (infection) & hospitalization (severe disease) | Clinical |
| Link-Gelles3 (2023) (42) | >=18 years of age | 2022-08-02 to 2022-09-21 | US | Documented | June 19 to August 20,2022 | Molecular test (primarily reverse transcription–polymerase chain reaction [RT-PCR] assay) | 61-90 & 121-180 | Documented | mRNA | Omicron (BA.4/BA.5) and afterwards | Emergency department and urgent care encounters (infection) & hospitalization (severe disease) | Clinical |
| Liu (2023) (43) | >=12 years of age | 2021-09-15 to 2021-10-04 | China | Documented | Between September 15, 2021 and October 4, 2021 | RT‐PCR | 0-30 & 31-60 & 61-90 | NA | Inactivated virus vaccines | Late-Delta | Infection (infection) & severe/critical (severe disease) | Non-clinical |
| Lutz (2023) (44) | >=12 years of age | 2021-01-01 to 2021-11-30 | US | Self-reported plus documented | From January 1, 2021, through November 30, 2021 | Molecular testing (reverse transcription polymerase chain reaction [PCR] or nucleic acid amplification test [NAAT]) or antigen test | 61-90 & 181+ | Testing | mRNA | Pre-Delta and Delta & late-Delta | Outpatient (infection) & inpatient (severe disease) | Non-clinical |
| Maeda2 (2023) (45) | >=16 years of age | 2022-01-01 to 2022-06-26 & 2021-07-01 to 2021-09-30 & 2022-01-01 to 2022-03-13 & 2022-04-11 to 2022-06-26 | Japan | Self-reported or documented | Between 1 January 2022, and 26 June 2022 | Nucleic acid amplification test, including polymerase chain reaction (PCR), loop-medical isothermal amplification (LAMP), nicking endonuclease amplification reaction (NEAR), and transcription-mediated amplification (TMA); and antigen quantification test | 31-60 & 91-120 & 121-180 & 181+ | NA | mRNA | Omicron (BA.1/BA.2) & late-Delta | Symptomatic (infection) | Clinical |
| Nasreen2 (2022) (46) | >=18 years of age | 2020-12-14 to 2021-09-30 | Canada | Documented | Same as study period, between December 2020 and September 2021 | Whole genome sequencing or screening PCR tests | 31-60 & 91-120 & 121-180 & 181+ | NA | mRNA & Adenovirus vector vaccines | Pre-Delta and Delta | Hospitalization or death (severe disease) | Clinical |
| Ng (2023) (47) | >=3 years of age | 2022-01-01 to 2022-05-31 | China | Documented | Same as study period, from 1 January to 31 May 2022 | RT-PCR | 91-120 & 181+ | RT-PCR or rapid antigen test | mRNA & Inactivated virus vaccines | Omicron (BA.1/BA.2) | Infection (infection) | Non-clinical |
| Powell (2022) (48) | 12-17 years of age | 2021-09-13 to 2022-01-12 | UK | Documented | 18/01/2022 to 31/12/2021 | PCR | 31-60 & 61-90 & 121-180 | Positive test result | mRNA | Late-Delta & Omicron (BA.1/BA.2) | Symptomatic (infection) | Clinical |
| Powell2 (2023) (49) | 12-17 years of age | 2021-08-09 to 2022-03-31 | UK | Documented | Between Aug 9, 2021, and March 31, 2022 | PCR | 61-90 & 181+ | PCR | mRNA | Late-Delta & Omicron (BA.1/BA.2) | Infection (infection) | Clinical |
| Price (2022) (50) | 12-18 years of age | 2021-07-01 to 2021-12-18 & 2021-12-19 to 2022-02-17 | US | Self-reported plus documented | Same as study period, from July 1, 2021, to February 17, 2022 | RT-PCR or antigen test | 31-60 & 91-120 & 181+ | NA | mRNA | Late-Delta & Omicron (BA.1/BA.2) | Hospitalization (severe disease) | Clinical |
| Prunas (2023) (51) | 12-16 years of age | 2021-06-15 to 2021-12-08 | Israel | Documented | Same as study period, from 15 June 2021, to 8 December 2021 | PCR | 31-60 & 121-180 & 181+ | Documented | mRNA | Late-Delta | Infection (infection) | Non-clinical |
| Qassim (2023) (52) | >=0 years of age | 2021-02-01 to 2022-11-30 | Qatar | Documented | Same as study period, between July 1, 2020 and November 30, 2022 | PCR | 31-60 & 61-90 & 121-180 & 181+ | PCR | mRNA | Pre-Delta and Delta & Omicron (BA.1/BA.2) & Omicron (BA.4/BA.5) and afterwards | Infection (infection) & severe, critical, or fatal COVID-19 (severe disease) | Non-clinical |
| Ranzani3 (2022) (53) | >=18 years of age | 2021-09-06 to 2021-12-14 & 2021-12-25 to 2022-04-22 | Brazil | Documented | December 25, 2021 to April 22, 2022 and September 6, 2021, to December 14, 2021 | RT-PCR or antigen detection test | 31-60 & 121-180 & 181+ | RT-PCR or antigen detection test | Inactivated virus vaccines | Late-Delta & Omicron (BA.1/BA.2) | Symptomatic (infection) & hospitalization or death (severe disease) | Clinical |
| Roberts (2022) (54) | >=18 years of age | 2021-01-01 to 2021-12-31 & 2021-01-01 to 2021-03-31 & 2021-04-01 to 2021-06-30 & 2021-07-01 to 2021-09-30 & 2021-10-01 to 2021-12-31 | US | Documented | Between January 1 and December 31, 2021 | RT-PCR | 61-90 & 91-120 & 121-180 & 181+ | Documented | mRNA | Late-Delta & pre-Delta and Delta | Infection (infection) & severe (severe disease) | Clinical |
| Rosa (2023) (55) | >=12 years of age | 2021-11-03 to 2022-06-20 & 2021-11-03 to 2021-11-30 | Brazil | Self-reported plus documented | Between November 3, 2021 and June 20, 2022 | RT-PCR | 0-30 & 31-60 & 91-120 & 121-180 & 181+ | NA | mRNA | Late-Delta & Omicron (BA.1/BA.2) | Symptomatic (infection) | Clinical |
| Self (2021) (56) | >=18 years of age | 2021-03-11 to 2021-08-15 | US | Self-reported plus documented | NA | RT-PCR or antigen test | 61-90 & 91-120 & 181+ | Self-reported | mRNA & Adenovirus vector vaccines | Pre-Delta and Delta | Hospitalization (severe disease) | Clinical |
| Skowronski (2022) (57) | >=18 years of age | 2021-05-30 to 2021-11-27 | Canada | Documented | Between epi-weeks 22-47 | NAAT | 0-30 & 31-60 & 91-120 & 121-180 & 181+ | NA | mRNA & Adenovirus vector vaccines | Late-Delta & pre-Delta and Delta | Infection (infection) & hospitalization (severe disease) | Non-clinical |
| Sritipsukho2 (2023) (58) | >=18 years of age | 2022-01-01 to 2022-06-15 | Thailand | Documented | 1 January 2022 to 15 June 2022 | RT-PCR | 61-90 & 91-120 & 121-180 | Documented | mRNA & Inactivated virus vaccines | Omicron (BA.1/BA.2) | Infection (infection) & moderate to critical diseases (severe disease) | Clinical |
| Stowe (2022) (59) | >=18 years of age | 2021-11-22 to 2022-02-02 | UK | Documented | From 25 November 2020 to 10 March 2022 | PCR | 61-90 & 91-120 & 121-180 | Testing | mRNA & Adenovirus vector vaccines | Omicron (BA.1/BA.2) | Hospitalization (severe disease) | Non-clinical |
| Suarez (2022) (60) | >=50 years of age | 2021-01-01 to 2021-12-12 | France | Documented | January 1st to December 12, 2021 | RT-PCR | 0-30 & 61-90 & 91-120 & 121-180 & 181+ | NA | mRNA | Pre-Delta and Delta & late-Delta | Symptomatic (infection) & hospitalization (severe disease) | Clinical |
| Suphanchaimat (2022) (61) | >=0 years of age | 2021-09-01 to 2022-12-31 | Thailand | Documented | Between 1 September 2021 and 31 December 2021 | PCR | 0-30 & 31-60 & 91-120 & 181+ | NA | Adenovirus vector vaccines & Inactivated virus vaccines | Late-Delta | Infection (infection) & severe (severe disease) | Non-clinical |
| Surie1 (2022) (62) | >=18 years of age | 2021-12-26 to 2022-06-19 & 2022-06-20 to 2022-08-31 | US | Self-reported plus documented | Same as study period, during December 26, 2021–August 31, 2022 | RT-PCR | 31-60 & 61-90 & 91-120 & 121-180 & 181+ | NA | mRNA | Omicron (BA.1/BA.2) & Omicron (BA.4/BA.5) and afterwards | Hospitalization (severe disease) | Clinical |
| Tabak (2021) (63) | >=18 years of age | 2021-05-01 to 2021-08-07 | US | Self-reported | Same as study period, May 1 to August 7, 2021 | PCR | 0-30 & 31-60 & 91-120 & 121-180 & 181+ | NA | mRNA & Adenovirus vector vaccines | Pre-Delta and Delta | Symptomatic (infection) | Clinical |
| Tamada (2023) (64) | >=20 years of age | 2021-07-01 to 2021-09-30 | Japan | Documented | Between July 1 and September 30, 2021 | PCR tests or antigen tests | 31-60 & 61-90 & 91-120 | Testing | mRNA | Late-Delta | Infection (infection) | Clinical |
| Tamandjou (2023) (65) | >=50 years of age | 2021-06-06 to 2022-02-10 | France | Documented | Within the period of June 6, 2021 and February 10, 2022 | RT-PCR | 0-30 & 61-90 & 91-120 & 121-180 & 181+ | Documented | mRNA | Late-Delta & Omicron (BA.1/BA.2) | Symptomatic (infection) & hospitalization (severe disease) | Clinical |
| Tan (2023) (66) | >=18 years of age | 2021-09-01 to 2022-04-25 | Singapore | Documented | From 1 September, 2021, to 30 November, 2021, and 1 December, 2021, to 25 April, 2022 | PCR | 0-30 & 91-120 & 181+ | NA | mRNA | Late-Delta & Omicron (BA.1/BA.2) | Infection (infection) & severe (severe disease) | Non-clinical |
| Tartof (2022) (67) | >=18 years of age | 2021-12-01 to 2022-02-06 | US | Documented | Same as study period, Dec 1, 2021, to Feb 6, 2022 | PCR | 31-60 & 121-180 & 181+ | PCR | mRNA | Late-Delta & Omicron (BA.1/BA.2) | Emergency department and urgent care encounters (infection) & hospitalization (severe disease) | Clinical |
| Tartof2 (2023) (68) | 5-11 years of age | 2021-11-01 to 2022-09-02 | US | Documented | Same as study period, from November 1, 2021 through September 2, 2022 | RT-PCR | 61-90 & 181+ | Laboratory confirmed | mRNA | Late-Delta & Omicron (BA.1/BA.2) | Emergency department and urgent care encounters (infection) | Clinical |
| Tartof3 (2022) (69) | 12-17 years of age | 2021-11-01 to 2022-03-18 | US | Documented | Same as study period, from November 1,2021, through March 18, 2022 | RT-PCR | 31-60 & 61-90 & 91-120 & 181+ | Documented | mRNA | Late-Delta & Omicron (BA.1/BA.2) | Emergency department and urgent care encounters (infection) | Clinical |
| Tartof4 (2022) (70) | >=18 years of age | 2022-05-09 to 2022-08-26 | US | Documented | Between May 9 and August 26, 2022 | PCR | 61-90 & 91-120 & 181+ | NA | mRNA | Omicron (BA.4/BA.5) and afterwards | Outpatient (infection) & hospitalization (severe disease) | Clinical |
| Tartof5 (2023) (71) | >=18 years of age | 2021-12-27 to 2022-06-04 | US | Documented | Dec 27, 2021 to June 4, 2022 | RT-PCR | 91-120 & 181+ | PCR | mRNA | Omicron (BA.1/BA.2) | Emergency department and urgent care encounters (infection) & hospitalization (severe disease) | Clinical |
| Tenforde3 (2022) (72) | >=18 years of age | 2021-03-11 to 2021-12-15 | US | Self-reported plus documented | NA | RT-PCR | 91-120 & 181+ | Laboratory confirmed | mRNA | Late-Delta | Hospitalization (severe disease) | Clinical |
| Thompson (2021) (73) | >=50 years of age | 2021-01-01 to 2021-06-22 | US | Documented | Same as study period, from January 1 through June 22, 2021 | Molecular testing | 0-30 & 31-60 & 61-90 & 91-120 & 121-180 & 181+ | Molecular or antigen assays | mRNA & Adenovirus vector vaccines | Pre-Delta and Delta | Emergency department and urgent care encounters (infection) & hospitalization (severe disease) | Clinical |
| Thompson2 (2022) (74) | >=18 years of age | 2021-08-26 to 2022-01-02 | US | Documented | Same as study period, August 26, 2021 to January 5, 2022 | Molecular testing (primar ily reverse transcription–polymerase chain reaction assay) | 91-120 & 181+ | NA | mRNA | Late-Delta & Omicron (BA.1/BA.2) | Emergency department and urgent care encounters (infection) & hospitalization (severe disease) | Clinical |
| Tseng (2022) (75) | >=18 years of age | 2021-12-06 to 2021-12-31 | US | Self-reported plus documented | Between 6 December 2021 and 31 December 2021 | Molecular diagnostic testing for SARS-CoV-2 | 0-30 & 31-60 & 181+ | COVID-19 diagnosis code or positive SARS-CoV-2 tests | mRNA | Late-Delta & Omicron (BA.1/BA.2) | Infection (infection) & hospitalization (severe disease) | Non-clinical |
| van Ewijk (2023) (76) | >=18 years of age | 2021-07-04 to 2021-12-08 | Netherlands | Self-reported | Between 4 July 2021 and 8 December 2021 | Lateral-flow antigen test (LFAT), a reverse transcription PCR (RT-PCR) test or a loop-mediated iso thermal amplification (LAMP) test | 61-90 & 91-120 & 181+ | Laboratory confirmed | mRNA & Adenovirus vector vaccines | Late-Delta | Infection (infection) | Non-clinical |

Table S3. Details of definitions of variables of included studies in the systematic review and meta-analysis.

| **Author (year)** | **Cases definition** | **Controls definition** | **Inclusion criteria** | **Exclusion criteria** | **Outcomes definition** | **Adjustments in analysis** |
| --- | --- | --- | --- | --- | --- | --- |
| Abu-Raddad1 (2022) (1) | Persons with a positive PCR test for SARS-CoV-2 | PCR-negative persons | Resident population of Qatar with PCR test | Received mixed vaccines, or who received a vaccine other than mRNA-1273, or who were tested by PCR after receiving a booster dose | Documented infection (a PCR-positive swab, regardless of the presence of symptoms) and against any severe Covid-19 cases (acute care hospitalizations), critical Covid-19 cases (intensive care unit hospitalizations), or fatal Covid-19 cases (Covid-19–related deaths) | Prior infection, healthcare worker status |
| Accorsi1 (2022) (2) | Those with positive SARS-CoV-2 NAAT | Those with negative SARS-CoV-2 NAAT | Adults 18 years of age or older who reported vaccination status (product and month and year of receipt of each dose) and at least one Covid-19 like symptom, which most likely reflected mild disease | Tests from persons who reported previous SARS-CoV-2 infection, or indeterminate test results, missing assay type, reported an immunocompromising condition (as COVID-19 vaccine recommendations differ for these individuals), missing data on sex or testing site census tract SVI, unknown vaccination status, receipt of a vaccination regimen other than the regimens of interest, receipt of the last vaccine dose within two weeks of the test date, vaccination before the month of the ACIP recommendation for primary or booster dose, receipt of a booster dose <2 or <5 months after the primary series for Ad26.COV2.S and mRNA primary series, respectively, or inconsistent vaccination information (e.g., reported receipt of vaccine but missing dose dates, reported no vaccine receipt but vaccine doses reported). Tests from participants with <2 weeks between the date of the last dose and the date of testing. Tests from participants with <2 weeks between the date of the last dose and the date of testing | Symptomatic SARS-CoV-2 infection determined by positive NAAT in a person reporting COVID-19-like illness | The number of days between the start of the analysis period and the test date, age group, sex, race, ethnic group, testing site location, Social Vulnerability Index of the U.S. census tract containing the testing site, and number of underlying chronic conditions |
| Andrejko2 (2023) (3) | Individuals testing positive for SARS-CoV-2 | Individuals testing negative for SARS-CoV-2 | California residents with SARS-CoV-2 molecular diagnostic test results | Participants aged ≤12 years, who were ineligible to receive vaccination until late in the study period, as well as participants who reported receiving vaccines other than BNT162b2 (Pfizer/BioNTech, New York, New York) or mRNA-1273 (Moderna, Cambridge, Massachusetts), due to limited observations | Symptomatic SARS-CoV-2 infection, defined as a positive SARS-CoV-2 test result with ≥1 symptom reported up to 14 days before testing | Age group, sex, region, and week of SARS-CoV-2 testing |
| Andrews1 (2022) (4) | Being due to the delta or omicron variant on the basis of whole-genome sequencing, genotyping, or S target status, with sequencing taking priority, followed by genotyping | Testing negative | Persons 18 years of age or older | Persons with four or more doses of vaccine, a heterologous primary schedule, or fewer than 19 days between their first dose and second dose | Symptomatic Covid-19 |  |
| Arashiro1 (2023) (5) | PCR-positive individuals | PCR-negative individuals | All individuals aged ≥16 years | Individuals who did not or could not consent to participate in the study, required immediate lifesaving treatment, or had previously participated in this study, individuals who had unknown symptom onset time, were tested ≥15 days after symptom onset, received vaccine types other than mRNA vaccines, or received unknown vaccine types | Symptomatic severe acute respiratory syndrome coronavirus 2 (SARS-CoV-2) infection | Age group, sex, presence of comorbidities, occupation (healthcare worker or not), SARS-CoV-2 diagnostic test in the past month, past SARS-CoV-2 infection, history of close contact, healthcare facility, calendar week, mask wearing, high-risk behavior, and influenza vaccination status for the 2022-2023 season |
| Arashiro2 (2022) (6) | PCR-positive individuals | PCR-negative individuals | All symptomatic individuals aged ≥20 years | Individuals who did not or could not consent to participate in the study, individuals who required immediate lifesaving treatment, and individuals who had previously participated in this study, individuals who had unknown symptom onset, were tested ≥15 days after symptom onset, or were tested during the nonepidemic period | Symptomatic severe acute respiratory syndrome coronavirus 2 infection | Age group, sex, presence of comorbidities, educational attainment, place of residence, occupation, SARS-CoV-2 diagnostic test in the past month, past SARS-CoV-2 infection, history of close contact, healthcare facility, and calendar week |
| Belayachi (2022) (7) | Subjects with severe or critical SARS-CoV-2 infection confirmed by the rt-PCR positive test result based on laboratory | Individuals who were tested negative for SARS-CoV-2 infection on an rt-PCR test; under the same conditions as the cases; based on laboratory data | Adults aged 18 years and above who had a reverse transcription real-time polymerase chain reaction (rt-PCR) test for SARS- CoV-2 infection | Recipients of BNT162b2 (Pfizer-BioNTech), Ad26. COV2. (Johnson & Johnson-Janssen), the ChAdOx1 nCoV-19 (AZD1222; Oxford-AstraZeneca, and those for whom 1-13 days had elapsed since receipt of the first dose Sinopharm COVID-19 vaccine | Vaccine effectiveness any time after receipt of the second dose and vaccine effectiveness by range time overtime after second dose receipt: (a) From 1st day to 30th days (during the first month after the second dose); (b) From 31th to 60th days (during the second month after the second dose); (C) From 61th to 90th days (during the third month after the second dose); (d) From 91st to120th days; (e) From 121th to 150th; and (f) beyond 150th day to 9th month | Sex, age, calendar days of the rt-PCR test; geographic location; and the 7-day moving average of the percentage of SARS-CoV-2 positive test |
| Brazete (2023) (8) | SARS-CoV-2 test-positive | SARS-CoV-2 test-negative | Individuals aged 18 years who were residents of Alto Minho, had at least one symptom included in the World Health Organization (WHO) COVID-19 definition, sought health care in a public emergency department in the region between 1 November 2021 and 2 March 2022, and were tested for SARS-CoV-2 using respiratory samples | Individuals who were not eligible for vaccination against COVID-19, those with unavailable laboratory test results, those without information on vaccination status and those with a symptom onset of more than 10 days before the test date. All individuals who had previously tested positive for COVID-19 were excluded from the analysis to minimise bias caused by natural immunity | Symptomatic SARS-CoV-2 infection confirmed with rRT-PCR tests, antigen tests or Xpress RT-PCR tests performed on respiratory samples from the nasopharynx or oropharynx; and moderate-to-severe disease associated with SARS-CoV-2 infection defined by hospitalisation over 24 h, intermediate or intensive care unit (I/ICU) admission or death with a recent positive test result | Age and the group of municipalities of residence |
| Buchan (2022) (9) | PCR-positive individuals | PCR-negative individuals | Individuals in Ontario whohadCOVID-19 symptoms, were aged 18 years or older, had provincial health insurance, and had a SARS-CoV-2 real-time reverse transcription-polymerase chain reaction (PCR) test between December 6 and 26, 2021 | Long-term care residents, individuals who had received only 1 dose or 4 doses of COVID-19 vaccine or who had received a second dose less than 7 days before being tested, individuals who tested positive for SARS-CoV-2 within the previous 90 days, individuals who had received 2 or 3 doses of ChAdOx1 vaccine(AstraZeneca) because estimated VE for that primary schedule is known to be lower and the product has rarely been used as a third dose, individuals who had received a vaccine not authorized by Health Canada for any dose, and individuals who had received the Ad.26.COV2.S vaccine | Individuals with confirmed SARS-CoV-2 infection using reportable disease and laboratory data | Age, sex, public health unit region of residence, number of SARS-CoV-2 polymerase chain reaction tests during the 3 months before December 14, 2020, SARS-CoV-2 infection more than 90 days before the index date, comorbidities, influenza vaccination status during the 2019-2020 and/or 2020-2021 influenza seasons, and neighborhood-level information on median household income, proportion of the working population employed as nonhealth essential workers, mean number of persons per dwelling, and proportion of the population who self-identified as belonging to a visible minority group |
| Buchan2 (2022) (10) | Those who test-positive | Those who test-negative | Individuals aged 12 to 17 years and SARS-CoV-2 tested (by real-time reverse transcription polymerase chain reaction) between November 22, 2021 (date of first Omicron detection) and March 6, 2022 using methods detailed elsewhere | NA | Symptomatic infection and severe outcomes (ie, hospitalization or death) | Age, sex, public health unit region of residence, comorbidities, influenza vaccination status during the 2019/2020 and/or 2020/2021 influenza seasons, positive test result >90 d before index date, week of testing, and neighborhood-level information on median household income, proportion of the working population employed as nonhealth essential workers, mean number of persons per dwelling, and proportion of the population who self-identify as a visible minority |
| Carazo2 (2023) (11) | Patients who were hospitalised for COVID-19 within 14 days after testing positive | Patients who tested negative | Older adults (aged ≥60 years) with symptoms associated with COVID-19 who were tested for SARS-CoV-2 in acute-care hospitals | Specimens from individuals who lived in long-term care facilities (because their baseline characteristics and hospital referral indications largely differed from the general population), those with missing comorbidity data, those with documented reinfection before the study period, those who received more than five vaccine doses or a non-mRNA vaccine dose, those vaccinated outside provincial minimum dosing intervals (specified as 21 days between first and second doses and 90 days between subsequent booster doses), those who were tested less than 14 days after their first vaccine or less than 7 days after any subsequent vaccine dose, those who had specimens collected less than 60 days after a positive result (according to reinfection definition), and those with negative specimens collected within 7 days before a positive result | Vaccine effectiveness against hospitalisation associated with BA.1, BA.2, and BA.4/5 infection | Sex, age, origin, epidemiological week, multimorbidity, chronic respiratory disease, chronic heart disease, cancer, obesity, immunosuppressive condition, neurological disease |
| Cerqueira-Silva (2022) (12) | Adults with a positive SARS-CoV-2 RT–PCR test | Adults with a negative SARS-CoV-2 RT–PCR test | All individuals aged 18 years or older who reported COVID-19-like symptoms and were tested for SARS-CoV-2 between 18 January 2021 and 11 November 2021 | (1) individuals younger than 18 years; (2) individuals who received a different vaccine for the second dose from the first; (3) individuals whose time interval between the first and second doses was fewer than 14 d; (4) tests with missing information of age, sex, city of residence or sample collection date; (5) negative test within 14 d of a previous negative test; (6) negative test followed by a positive test up to 7 d; (7) any test after a positive test up to 90 d; and (8) tests with a symptom onset date greater than notification date | Cases of COVID-19 hospitalization or death (a positive SARS-CoV-2 test accompanied by hospitalization or death occurring within 28 d of the sample collection date). Controls for the outcome of hospitalization or death were defined based on a negative test |  |
| Cerqueira-Silva3 (2022) (13) | Adults with a positive SARS-CoV-2 RT–PCR test | Controls with a negative SARS-CoV-2 RT–PCR test | All individuals aged 18 years or older who reported COVID-19-like symptoms and were tested for SARS-CoV-2 between 18 January 2021 and 11 November 2021 | (1) individuals younger than 18 years; (2) individuals who received a different vaccine for the second dose from the first; (3) individuals whose time interval between the first and second doses was fewer than 14 d; (4) tests with missing information of age, sex, city of residence or sample collection date; (5) negative test within 14 d of a previous negative test; (6) negative test followed by a positive test up to 7 d; (7) any test after a positive test up to 90 d; and (8) tests with a symptom onset date greater than notification date | Infection and severe COVID-19 outcomes (hospitalization or death) | Age, sex, temporal trends, state of residence, previous infection, pregnancy, postpartum period and comorbidities |
| Cerqueira-Silva4 (2022) (14) | Individuals with RT-PCR / Lateral-flow test positive | Individuals with RT-PCR / Lateral-flow test negative | All individuals aged 18 years or older who reported COVID-19-like symptoms and were tested for SARS-CoV-2 between January 01, 2022, and April 17, 2022 | The exclusion criteria for tests were: (i) tests from individuals younger than 18 years; (ii) tests from individuals who received a different vaccine for the second dose from the first; (iii) tests from individuals whose time interval between the first and second doses was less than 14 days; (iv) tests from individuals with less than 115 days between the second and booster dose (outside the interval between doses officially recommended in Brazil); (v) tests with missing information of age, sex, city of residence or sample collection date; (vi) negative tests from individuals with a positive test; (vii) more than three doses of COVID-19 vaccines | Symptomatic infection and severe outcomes (hospitalization or death) | Age, sex, temporal trends, state of residence, previous infection, municipality deprivation index, and comorbidities |
| Chatzilena (2023) (15) | Patients admitted with aLRTD and a positive admission SARS-CoV-2 test, using the UKHSA diagnostic assay in current use | Controls had to have aLRTD and a negative SARS CoV-2 result | Patients who had signs/symptoms of respiratory infection and were aged ≥18 y (years) on hospitalisation | SARS-CoV-2 positive admissions without identified variant and SARS-CoV-2 negative admissions between 8th Nov 2021 and 6th Feb 2022 | VE against hospital admission with either a clinical or radiological aLRTD diagnosis or aLRTD signs/symptoms was assessed | Age, gender, index of multiple deprivation, Charlson comorbidity index, time, and community infection prevalence |
| Chemaitelly1 (2021) (16) | PCR-positive persons | PCR-negative persons | Resident population of Qatar | All persons who received mixed vaccines, or who received a vaccine other than BNT162b2 | Documented infection (a PCR-positive swab, regardless of the reason for PCR testing or the presence of symptoms),severe(a severe acute respiratory syndrome coronavirus 2 (SARS-CoV-2) infected person with “oxygen saturation of <90% on room air, and/or respiratory rate of >30 breaths/minute in adults and children >5 years old (or ≥60 breaths/minute in children <2 months old or ≥50 breaths/minute in children 2-11 months old or ≥40 breaths/minute in children 1-5 years old), and/or signs of severe respiratory distress (accessory muscle use and inability to complete full sentences, and, in children, very severe chest wall indrawing, grunting, central cyanosis, or presence of any other general danger signs)), critical(a SARS-CoV-2 infected person with “acute respiratory distress syndrome, sepsis, septic shock, or other conditions that would normally require the provision of life sustaining therapies such as mechanical ventilation (invasive or non-invasive) or vasopressor therapy),or fatal case (a death resulting from a clinically compatible illness, in a probable or confirmed COVID-19 case, unless there is a clear alternative cause of death that cannot be related to COVID-19 disease (e.g. trauma)) of Covid-19 | Previous infection and health care worker status |
| Chemaitelly3 (2022) (17) | Persons infected with BA.1, BA.2, or any-Omicron-sub-variant | Uninfected persons | Resident population of Qatar | All persons who received a vaccine other than BNT162b2 or mRNA-1273, or who received a different mix of vaccines | Symptomatic SARS-CoV-2 Omicron (B.1.1.529)1 infection | Sex, 10-year-age group, nationality, and calendar week of PCR test |
| Chung3 (2022) (18) | Individuals who tested positive at least once during the study period | Those who tested negative throughout | Ontario residents who were aged ≥16 years, registered for provincial health insurance, and not residing in a long-term care facility as of December 14, 2020 | Who had previously tested positive for SARS-CoV-2 or had received only a single vaccine dose, had received 2 doses but were <7 days from the second dose, had received any non–Health Canada–approved vaccines (including the Johnson & Johnson/Janssen Ad26.COV2.S vaccine, which was approved but rarely used in Ontario), or had received 3 doses | VE against any infection, symptomatic infection, and severe outcomes (COVID-19-associated hospitalizations or death) separately | Age, sex, public health unit region, biweekly period of test, number of SARS-CoV-2 tests in the 3 months before December 14, 2020, presence of any comorbidity that increases the risk of severe COVID-19, receipt of influenza vaccination in the current or prior influenza season, and neighborhood-level household income, persons per dwelling, proportion of persons employed as nonhealth essential workers, and self-identified visible minority quintiles |
| Ciesla (2023) (19) | COVID-19-like illness symptoms with a positive NAAT result | COVID-19-like illness symptoms with a negative NAAT result | Persons aged ≥5 years with ≥1 coronavirus disease-2019 (COVID-19)-like symptoms and a SARS-CoV-2 nucleic acid amplification test from April 2 to August 31, 2022 | Immunocompromising condition, or individuals reporting any prior SARS-CoV-2 infection, or aged 5 to 49 years and received >3 doses or were aged ≥50 years and received >4 doses, or received a booster dose earlier (<5 months) than the recommended interval, or received any doses of a non-mRNA vaccine, or received only 1 dose of vaccine, or were missing vaccination information or covariables of interest | Symptomatic SARS-CoV-2 infection | Calendar day of test, age, gender, race, ethnicity, 2018 CDC census tract social vulnerability index (SVI), underlying conditions, individual state of residence, pharmacy chain, and average cases per 100,000 by site zip code |
| Collie2 (2022) (20) | Tested positive | Tested negative | Patients that had been hospitalized for medical treatment in South Africa from the 15 November 2021 to the 24 June 2022 | COVID19 PCR test results associated with admissions unlikely to be related to COVID19 treatment. Indeterminate test results, test results for individuals younger than age 18, or test results for individuals who joined the medical scheme over the study period (i.e. individuals with unknown prior vaccination history), or test results for individuals vaccinated with a single dose of Pfizer or a dose of J&J | In-hospital admission | Age, sex, number of documented CDC risk factors, surveillance week, period of prior documented infection, and geographic region |
| Ella (2023) (21) | Those who tested positive for SARS-CoV-2 | Those who tested negative | KPSC members ≥18 years of age between May 9 and August 26, 2022 who were diagnosed with acute respiratory infection and tested for SARS-CoV-2 on polymerase chain reaction in one of four healthcare settings including outpatient visits (which included virtual appointments), urgent care centers, emergency departments (ED), or the hospital | Individuals who received a COVID-19 vaccine other than BNT162b2, who received only one dose of BNT162b2, or received >3 doses | omicron subvariants BA.4 and BA.5, by highest level of care | Week of COVID-19 health-care encounter, age, sex, race or ethnicity, previous SARS-CoV-2 infection, BMI, Charlson score, and history of previous influenza and pneumococcal vaccination, and nirmatrelvir plus ritonavir receipt |
| Embi2 (2023) (22) | Tested positive | Tested negative | Patients were aged ≥18 years with ≥1 discharge diagnosis among a previously published list of qualifying COVID-19–like illness (CLI) diagnoses | mRNA vaccine recipients who received 1) only 1 dose, 2) second dose 1–13 days earlier, 3) third dose 1–6 days earlier, or 4) ≥4 doses. Those who received 1) first dose 1–13 days earlier, 2) second dose (any product) 1–6 days earlier, or 3) ≥3 doses. Patients with a positive SARS-CoV-2 result 15–89 days earlier | laboratory-confirmed COVID-19-associated emergency department (ED) or urgent care (UC) events and hospitalizations | Age, geographic region, calendar time, prior positive SARS-CoV-2 test result, and local SARS-CoV-2 circulation on the day of each medical visit, and weighted for patients' inverse propensity to be vaccinated or unvaccinated |
| Ferdinands (2022) (23) | Those whose outcome was confirmed COVID-19 | Those with COVID-19–like illness and negative SARS-CoV-2 test results | Adults aged ≥18 years with a COVID-19–like illness diagnosis | Recipients of Janssen vaccine, 1 or >3 doses of an mRNA vaccine, and those for whom <14 days had elapsed since receipt of any dose | Medical encounters (adults aged ≥18 years with a COVID-19–like illness diagnosis who had received molecular testing for SARS-CoV-2, the virus that causes COVID-19, during the 14 days before through 72 hours after the medical encounter); hospitalization | Age, local virus circulation, propensity to be vaccinated, and other factors |
| Ferdinands2 (2022) (24) | Patients with covid-like illness with laboratory confirmed covid-19 | Patients with covid-like illness and negative SARS CoV-2 test results (controls could have had positive test results for other respiratory viruses such as influenza) | adults (≥18 years) who received care for covid-like illness at a VISION network hospital or emergency department or urgent care center and had molecular testing for SARS-CoV-2 at least 14 days after vaccines became locally available for their age group | Individuals who received any vaccine other than the BNT162b2 or mRNA-1273 vaccine, individuals who received more than four doses of an mRNA vaccine before the index medical contact, individuals who received only one dose of an mRNA vaccine less than 14 days before the index contact or who had a third or fourth dose less than seven days before the index contact, individuals known to have a positive laboratory test result for a SARS-CoV-2 infection more than 14 days before the index contact, and individuals with a positive SARS-CoV-2 test result but no diagnoses or symptoms suggesting covid-19 illness | Positive or negative molecular SARS-CoV-2 result for a test done within 14 days before a medical contact to less than 72 h after among patients presenting with covid-like illness, as identified from ICD-9 and ICD-10 diagnostic codes | Age, race, ethnicity, local virus circulation, immunocompromised status, and likelihood of being vaccinated |
| Fleming-Dutra (2022) (25) | Those with positive SARS-CoV-2 NAAT results | Those with negative NAAT results | Children and adolescents aged 5 to 15 years reporting 1 or more symptoms tested at the pharmacy chain from December 26, 2021, to February 21, 2022 | Indeterminate test results; missing assay type, reported an immunocompromising; unknown vaccination status, vaccine product other than BNT162b2; receipt of 1 vaccine dose or receipt of the second or third dose within 2 weeks of the test date; vaccination before the month of the recommendation by the Advisory Committee on Immunization Practices; receipt >2 dose for non-immunocompromised children 5-11 years or >3 dose for adolescents 12-15 years; receipt of a third dose < 4 months after the second dose for adolescents, or inconsistent vaccination information | Symptomatic SARS-CoV-2 infection (determined by positive NAAT result in a person reporting COVID-19–like illness) | Calendar day of test (continuous variable), race, ethnicity, sex, testing site region, and testing site census tract Social Vulnerability Index |
| Fleming-Dutra2 (2023) (26) | Those who received a positive SARS-CoV-2 test result | Those who received a negative test result | NAATs from children with one or more COVID-19–like illness symptom | The caregiver reported any of the following conditions: immunocompromise, positive SARS-CoV-2 test within 3 months, receipt of a non-mRNA COVID-19 vaccine or mixed product regimen, COVID-19 vaccine dose receipt within 2 weeks of test date, or third COVID-19 vaccine dose received during or after December 2022 |  | Single year of age, gender, race, ethnicity, Social Vulnerability Index of the testing location, underlying conditions, U.S. Department of Health and Human Services region of testing site, pharmacy chain conducting the test, local incidence, and testing calendar date |
| Florentino2 (2022) (27) | Individuals with a positive SARS-CoV-2 RT-PCR or antigen test | Individuals with a negative SARS-CoV-2 RT-PCR or antigen test | Adolescents aged 12–17 years with symptoms indicative of COVID-19 | Negative SARS-CoV-2 tests recorded within 14 days of a previous negative test, negative tests recorded within 7 days after a positive test, any test done within 90 days after a positive test, and tests with missing sex and location information | COVID-19 symptomatic infection, confirmed by rapid antigen testing or RT-PCR in Brazil and only by RT-PCR in Scotland | Age, sex, epidemiological week, state of residence, socioeconomic position measured by quintile of deprivation, previous SARS-CoV-2 infection, and number of comorbidities commonly associated with COVID-19 illness |
| Grewal2 (2023) (28) | COVID-19-associated hospitalization or death due to, or partially due to, COVID-19 | Controls had to be symptomatic and test negative for SARS-CoV-2, but may or may not have had asevere outcome | Community-dwelling adults aged ≥50 years who had ≥1 reverse-transcription polymerase chain reaction (RT-PCR) test for SARS-CoV-2 between January 2, 2022 and October 1, 2022 | Immunocompromised individuals or those who received a bivalent mRNA vaccine, Ad26.COV2, or >1 dose of ChAdOx1-S by the index date. Delta (B.1.617.2) cases identified using whole genome sequencing or based on an S-gene target positive screening result before January 24, 2022. Hospitalizations when specimen collection occurred >3 days after admission and those flagged as being nosocomial | Severe outcomes (hospitalization or death) | Sex, age, public health unit region, four area-level variables representing different socio-demographic characteristics, influenza vaccination during 2019/2020 or 2020/2021, SARSCoV-2 infection >90 days prior, number of SARS-CoV-2 tests within 3 months prior to December 14, 2020, comorbidities, receipt of home care services, and week of test |
| Heidarzadeh (2023) (29) | Patients admitted to hospitals in Guilan Province who had a positive PCR test | People visiting public health centers to get a COVID-19 diagnosis with negative PCR tests | Population aged 5 years and above | Unspecified PCR test results and having received mixed vaccines | Temporary admission: The presence of respiratory symptoms (dyspnea, pain, and pressure in the chest, etc.) with or without a fever, SpO 2 of 90-94%, lung involvement < 50%. Regular admission: respiratory rate > 30, SpO2 < 90%, lung involvement > 50% on the Computed Tomography scan. ICU admission: Symptoms of respiratory failure despite non-invasive oxygen therapy, symptoms of septic shock, multiple organ failure. Death: The occurrence of death in a person with a probable or definite diagnosis of COVID-19 that is clinically ruled to have been caused by COVID-19 without there being other specific non-COVID-19-related causes (e.g., trauma) and the absence of a full recovery period between active COVID-19 and death. | Age group, sex, week sampling polymerase chain reaction (PCR), health care workers, history of PCR positive |
| Husin (2022) (30) | Positive tests for SARS-CoV-2 infection (RT-PCR or RTK-Ag) | Negative tests | Adolescents aged 12 to 17 years in Malaysia | NA | SARS-CoV-2 infections | Age, sex, states of residence, strata (urban/rural), school types, and number of baseline (before September 1, 2021) tests |
| Ionescu (2023) (31) | 12–17-year-olds with NAAT-confirmed SARS-CoV-2 infection during the study period | NAAT negative | 12–17-year-olds in Quebec and British Columbia, Canada | Participants identified as having had a NAAT-confirmed SARS-CoV-2 infection before 5 September 2021. Those with missing information and vaccination that was invalid or involved <2 doses, 2 doses <21 days apart, 3 doses <24 weeks after the second, or with any vaccine other than BNT162b2. | SARS-CoV-2 infection | Age, sex, epi-week and region |
| Khan (2022) (32) | Testing positive for SARS-CoV-2 | Those testing negative | Children aged 5 to 11 years who were tested for SARS-CoV-2 via PCR at a Walgreens pharmacy between November 2, 2021, and September 30, 2022 | Who had received a COVID-19 vaccine other than BNT162b2, were vaccinated before the month of recommendation by the Advisory Committee on Immunization Practices (November2021), received a third dose less than 5 months after the second dose, or self-reported having an immunocompromising condition and who were tested between December 10, 2021, and January 15, 2022 | Confirmed SARS-CoV-2 infection (regardless of the presence of symptoms) | Age, gender, race/ethnicity, recent close contact with someone suspected or confirmed to have COVID-19, testing volume of pharmacy, rural/suburban/urban trade area, and calendar week |
| Kim2 (2022) (33) | Participants with a positive SARS-CoV-2 result | Participants with only negative SARS-CoV-2 results | Eligible participants who had fever, cough, or loss of taste or smell and sought outpatient medical care or clinical SARS-CoV-2 testing within 10days of illness onset. | Participants who self-reported COVID-19 vaccination but were missing verified documentation of doses received or receiving a non-mRNA vaccine. | SARS-CoV-2 infection | Age, site, illness onset week, and prior infection status |
| Klein (2023) (34) | Who had >=1 CLI code in any 1 of the 4 categories and >=1 positive severe acute respiratory syndrome coronavirus 2 (SARS-CoV-2) reverse transcription polymerase chain reaction (RT-PCR) test result | Who had >=1 CLI code and negative RT-PCR test results | All patients aged 5 to 17 years with eligible ED/UC encounters or hospitalizations from April 2021 through September 2022 | Patients vaccinated before the Advisory Committee on Immunization Practice recommendation date for their age, patients who received only 1 vaccine dose, or if index date was <14 days after receipt of dose 2 or <7 days after dose 3. Who received a third dose before monovalent boosters were recommended for their age, a monovalent booster <5 months after dose 2, >3 doses of vaccine, or mRNA-1273 vaccine (Moderna). Patients who were likely immunocompromised | ED/UC encounters and hospitalizations | Age, geographic region, and spline functions of calendar time, the time-and area-specific percent of SARS-CoV-2 tests that were positive, and a propensityto-be-vaccinated score |
| Klein1 (2022) (35) | Those who with acute laboratory-confirmed COVID-19 | Those who without acute COVID-19 | Persons aged ≥5 years with a COVID-19–like illness diagnosis who had received SARS-CoV-2 molecular testing | Patients were excluded if they 1) were vaccinated before the CDC recommendation date for their age group, 2) received a third dose before booster doses were recommended for their age group, 3) received a booster dose 3 doses of the vaccine, or 5) if <14 days had elapsed since receipt of dose 2 or <7 days since dose 3. Patients who were likely immunocompromised based on diagnosis codes were also excluded | COVID-19-associated emergency department and urgent care encounters and hospitalizations | Age, geographic region, calendar time, and local virus circulation |
| Lewis2 (2022) (36) | COVID-19–like illness and received a positive SARS-CoV-2 test result (nucleic acid amplification test [NAAT] or antigen test) within 10 days of illness onset | Test-negative controls (hospitalized with signs or symptoms of an acute respiratory illness but testing negative for SARS-CoV-2 by NAAT) or syndrome-negative (controls hospitalized without signs or symptoms of an acute respiratory illness and testing negative for SARS-CoV-2 by NAAT) | Adults who received a single dose Ad26.COV2.S vaccine ≥14 days before illness onset. Methods have been described in detail elsewhere | Participants who received other vaccine products (mRNA vaccine, or multiple doses of AD.26.COV2.S) | COVID-19–associated hospitalization | Admission date (biweekly intervals), geographic region, age group, sex, and self-reported race and Hispanic ethnicity |
| Lim (2022) (37) | Person with RT-PCR confirmation of infection with SARS-CoV-2 irrespective of clinical signs or symptoms. COVID-19 ICU admissions referred to confirmed COVID-19 cases that were admitted to ICU based on clinical judgement due to disease progression to severe disease, such as category 4 (requiring oxygen therapy) or category 5 (requiring mechanical ventilation) | Individuals with negative RT-PCR test and without a positive test in the previous 90 days or the following 14 days | Resident population of Labuan aged 18 years who had been tested for SARS-CoV-2 by RT-PCR between 1 March 2021 and 31 October 2021 | Individuals with previous infections or had received COVID-19 vaccine other than BNT162b2 vaccine. | COVID-19 infection ascertained by RT-PCR tests for SARS-CoV-2 on respiratory specimens, comprising of samples from the nasopharynx and saliva | Age, sex, nationality, reason for testing and testing dates |
| Lind1 (2023) (38) | Positive RT-PCRs | Negative RT-PCRs | vaccine-eligible people (aged ≥16 years) | People who received a vaccine prior to state distribution (14 December 2020), had a previous positive SARS-CoV-2 RT-PCR or rapid antigen test, or had missing covariate information, or tests performed after receiving a booster (third) dose or an Ad26.COV2 vaccine dose | WGS-classified Alpha-, Delta-, and other-variants infection, symptomatic infection, and COVID-19–associated hospitalizations | Date of test, age, sex, race/ethnicity, Charlson comorbidity score, number of nonemergent YNHH encounters in the year prior to vaccine rollout in Connecticut, insurance group (uninsured, Medicaid, Medicare, other), social vulnerability index of residential zip code (continuous), and residential county |
| Lind2 (2022) (39) | Positive tests | Negative tests | vaccine-eligible (>=5 years of age) people who were alive at the beginning of the study period and had at least one SARS-CoV-2 test in the electronic medical records (EMRs) | Tests that were performed after receiving a heterologous primary vaccination (i.e., different first and second dose manufacturers) or an Ad26.COV2 vaccine dose. Tests that were performed among people who received booster doses prior to eligibility (defined as five months since second primary vaccine dose and after booster vaccination approval in the US. Tests that were performed in the 90 days after a positive SARS-CoV-2 test (rapid antigen or RT-PCR), had a positive reflex result with an inconclusive SGTF finding, were obtained from people with more than one prior SARS-CoV-2 infection or with missing confounder data, or occurred after a second booster (fourth) dose. | SARS-CoV-2 infections | Date of test, age, sex, race/ethnicity, Charlson comorbidity score, number of nonemergent visits in the year prior to the vaccine rollout in Connecticut, insurance status, municipality, and SVI of residential zip code |
| Link-Gelles1 (2023) (40) | Those who received a positive SARS-CoV-2 test result | Those who received a negative test result | Children aged 6 months–5 years who received molecular SARS-CoV-2 testing during August 1, 2022–June 17, 2023 | Most recent vaccine dose was received <14 days before the index date, child had received a combination of Moderna and Pfizer-BioNTech vaccine doses, or a vaccination schedule that was not authorized in the study population had been used (e.g., 4 monovalent Pfizer BioNTech doses or 3 monovalent Moderna doses) | emergency department or urgent care (ED/UC) encounters | Age, sex, race and ethnicity, geographic region, and calendar time |
| Link-Gelles2 (2022) (41) | Persons with positive SARS-CoV-2 test results | Persons with negative SARS-CoV-2 test results, | Adults with COVID-19–like illness and a SARS-CoV-2 molecular test |  | 1) a medical event occurred during the washout period; 2) a likely immunocompromising condition was present; 3) an mRNA vaccine dose was received before it was recommended§§; 4) any doses of a non-mRNA vaccine such as JNJ-78436735 (Janssen [Johnson & Johnson]) were received; 5) <14 days had elapsed since receipt of dose 2 or <7 days since receipt of dose 3 or dose 4; or 6) a previous SARS-CoV-2 infection was documented in the patient’s medical record before the index encounter | Age, geographic region, calendar time, and local virus circulation and weighted for inverse propensity to be vaccinated or unvaccinated |
| Link-Gelles3 (2023) (42) | Patients with at least 1 COVID-19–like illness code and a positive SARS-CoV-2 molecular test result | Patients with at least 1 COVID-19–like illness code and a negative SARS CoV-2molecular test result | Adults aged 18 years or older with a medical encounter related to COVID-19–like illness and SARS-CoV-2 molecular testing | Those who received a third or fourth dose before recommended for immunocompetent adults or received a dose with a shorter interval than recommended (ie, less than 5 months between second and third dose or less than 4 months between third and fourth dose) or those who received only 1 mRNA vaccine dose, received a non-mRNA vaccine (eg, viral vector), or had a likely immunocompromising condition, as previously defined | COVID-19–like illness encounters include ICD-9 or ICD-10 codes for acute respiratory clinical diagnoses (eg, pneumonia, respiratory failure) or COVID-19–related signs or symptoms (eg, shortness of breath, cough, fever) during an UC or ED visit or a hospital admission with at least 24 hours’ duration. | Age, geographic region, calendar time, and local virus circulation and weighted for inverse propensity to be vaccinated or unvaccinated |
| Liu (2023) (43) | RT‐PCR positive | A person would be identified as a close contact, including who has close contact with confirmed and suspected cases in the past 2 days before the onset of symptoms, or who has close contact with asymptomatic infected persons in the past 2 days before sampling without effective protection | Populations aged 12 years or older | The population aged <12 years | The severe COVID‐19 was defined according to the guideline for the diagnosis and treatment of COVID‐19 (eighth trial revised version) | Gender and age |
| Lutz (2023) (44) | Those with a positive SARS-CoV-2 result within the 10 days before illness onset or 4 days following their visit/admission and ≥1 symptom consistent with COVID-19-like illness | Those with negative results and ≥1 symptom consistent with COVID-19-like illness | Enrolled in the Respiratory Surveillance in Native American Children and Adults Study, and eligible to receive any COVID-19 vaccine (based on age and enrollment site), and American Indian or Alaska Native person, and clinical (PCR, NAAT, antigen) and/or research (PCR only) result for SARS-CoV-2 testing obtained within 10 d of onset of the enrollment illness and within 4 d of presenting for care, and medically attended event (outpatient visit [outpatient clinic or emergency department] or hospital admission), and one of the following indicative of COVID-like illness: 1)at least one of the following: cough, shortness of breath, difficulty breathing, new olfactory disorder, new taste disorder; or 2)at least 2 of the following: fever, chills, rigors, myalgia, headache, sore throat, nausea or vomiting, diarrhea, fatigue, congestion or coryza; or 3)severe respiratory illness with at least one of the following: clinical or radiographic evidence of pneumonia or ARDS | Presented for care >14 d after illness onset, or first SARS-CoV-2 test associated with the illness occurred >10 d before illness onset, or first SARS-CoV-2 test associated with the illness occurred >4 d after presenting for care, or test results unavailable, or enrolled as a control and any positive test for acute SARS-CoV-2 infection in the 14 d before illness onset, or received the first dose of a 2-dose series 0–13 d before illness onset, or received a second dose of the Pfizer-BioNTech vaccine <17 d after the first or received a second dose of the Moderna vaccine <24 d after the first, or received a COVID-19 vaccine other than an mRNA product (i.e., Janssen), or received a booster dose (any product) >7 d before illness onset, or missing date for vaccination receipt, or unable to ascertain/missing vaccination status, or if positive for SARS-CoV-2, previous enrollment as a case in the study, or if negative for SARS-CoV-2, previous enrollment as a control in the study | Laboratory-confirmed COVID-19-associated outpatient visits (outpatient clinic and emergency department) and hospitalizations before and during Delta predominance | Age, sex, month of illness |
| Maeda2 (2023) (45) | An episode with an individual with signs or symptoms and a positive test result for SARS-CoV-2 | An episode with an individual with signs or symptoms but a negative result for SARS-CoV-2 | Individuals aged ≥16 visiting medical facilities with COVID-19-related signs or symptoms from 1 January to 26 June 2022 | Episodes involving individuals receiving COVID-19 vaccines other than BNA162b2 and mRNA-1273; results generated by the rapid antigen test; episodes involving an unknown vaccination status; episodes with no information on the vaccination date; episodes involving individuals with previous COVID-19 histories; episodes involving individuals with multiple episodes: (1) episodes with multiple negative test results and identical symptom onset date; (2) episodes with negative test results within seven days after a previous negative result; (3) episodes with negative test results within three weeks before or after a positive test result | Symptomatic SARS-CoV-2 infections | Age, sex, underlying medical conditions, calendar week of test, history of contact with COVID-19 patients within 14 days, healthcare professional status, and medical facilities |
| Nasreen2 (2022) (46) | Subjects with COVID-19 hospitalizations and deaths | Symptomatic subjects who tested negative during the study period | All residents aged ≥18 years, eligible for provincial health insurance, not living in long-term care, and tested for SARS-CoV-2 between the start of vaccine availability in a province (Ontario, Quebec: 14 December 2020; BC: 15 December; Manitoba: 16 December) and 30 September 2021 and met our case or control definitions | Recipients of non-Health Canada-authorized vaccines or Ad26.COV2.S (Johnson & Johnson Janssen) vaccine. Nosocomial cases flagged in notifiable disease reporting systems and SARS-CoV-2-positive cases with specimen collection >3 days after hospital admission. | COVID-19 hospitalization or death identified from notifiable disease reporting systems and/or other administrative databases. COVID-19 hospitalization was defined as hospitalization or ICU admission with a positive SARS-CoV-2 test within 14 days prior to or 3 days after hospitalization | Age group, sex, public health unit region, biweekly period of test, number of SARS-CoV-2 tests in the 3 months prior to 14 December 2020, presence of any comorbidity that increase the risk of severe COVID-19, receipt of influenza vaccination in current or prior influenza season, and neighbourhood income (material deprivation index in British Columbia), proportion of persons employed as non-health essential workers, persons per dwelling, and self-identified visible minority quintiles |
| Ng (2023) (47) | Subjects who had initial negative result(s) but became positive after admission | Eligible sex and age matched persons who tested negative during the same calendar month | Subjects who received a real-time reverse transcription polymerase chain reaction (RT-PCR) based COVID-19 test from the Prince of Wales Hospital | Cases or controls known to have been infected before, either based on RT-PCR or rapid antigen test; records without valid results, duplicate tests of the same specimen, tests for children under 3 years old, and patients without available medical record, previous history of COVID-19, or having concurrent major illnesses not related to COVID-19 precluding assessment on disease severity | SARS-CoV-2 infection of all severity ascertained by RT-PCR testing on respiratory specimens | Sex, age, calendar date of testing and Charlson Comorbidity Index |
| Powell (2022) (48) | The Delta or Omicron variant based on whole genome sequencing, genotyping or S-gene target status, with sequencing taking priority, followed by genotyping followed by SGTF status | Symptomatic adolescents in the same age groups who had a negative SARS-CoV-2 PCR test | Adolescents | Cases with booster doses or unusual vaccine schedules | PCR-confirmed symptomatic infection with the delta and omicron variants of SARS-COV-2 | Age, sex, index of multiple deprivation (quintile), ethnic group, geographic region (NHS region), period (calendar week of onset), clinical risk group status (a separate flag for those aged over and under 16), clinically extremely vulnerable (if aged 16 and above) and previous positivity |
| Powell2 (2023) (49) | PCR-confirmed COVID-19 | Participants who had a negative SARS-CoV-2 PCR test | Symptomatic adolescents aged 12-17 years who were unvaccinated, or had received primary BNT162b2 immunisation, at symptom onset and had a community (Pillar 2) SARS-CoV-2 PCR test | Negative tests taken within 7 days of a previous negative test, and negative tests where the symptom onset date was within the 10 days of a symptom onset date for a previous negative test, or negative tests taken within 21 days of a subsequent positive test, or positive and negative tests within 90 days of a previous positive test, or positive tests with no sequencing, genotyping, or SGTF between Nov 29, 2021, and Jan 4, 2022 | Protection against SARS-CoV-2 delta and omicron infection | Age, sex, index of multiple deprivation quintile, ethnic group, geographical region, period, clinical risk group status, and clinically extremely vulnerable |
| Price (2022) (50) | Hospitalized with Covid-19 as the primary reason for admission or with a clinical syndrome consistent with acute Covid-19 (≥1 of fever, cough, shortness of breath, loss of taste, loss of smell, gastrointestinal symptoms, receipt of respiratory support, or new pulmonary findings on chest imaging), and had a positive SARS-CoV-2 RT-PCR or antigen test result within 10 days after symptom onset or within 72 hours after hospital admission | Hospitalized patients with a negative SARS-CoV-2 RT-PCR or antigen test result, with or without Covid-19– associated symptoms | Case patients with Covid-19 and controls without Covid-19 at 31 hospitals in 23 states | Patients who received the SARS-CoV-2 test result more than 10 days after illness onset or more than 72 hours after the admission date, those who were partially vaccinated, those who were vaccinated 0-13 days before symptom onset, those whose vaccination status was unknown, and those who had received the mRNA-1273 (Moderna) or Ad26.COV2.S (Johnson & Johnson–Janssen) vaccine, neither of which was authorized for adolescents younger than 18 years of age during the study period. Patients admitted for reasons not related to Covid-19 (e.g., trauma or suicide attempt) who had a positive SARS-CoV-2 test during admission | Hospitalization | Sex, age, race, region, calendar time |
| Prunas (2023) (51) | Individuals with a positive PCR test occurring after 15 June 2021 | Individuals who had not tested positive prior to the date of the positive PCR of their matched case, that is, individuals who either had a negative PCR test result or who did not obtain a PCR test | MHS members, aged 12–16 years, who received either 1 or 2 doses of the BNT162b2 vaccine | Adolescents aged 17 years or older | SARS-CoV-2 infection (regardless of the presence of symptoms) and symptomatic SARS-CoV-2 infection (COVID-19) | Obesity and the number of positive tests performed on that day throughout the entire population |
| Qassim (2023) (52) | SARS-CoV-2-positive tests | SARS-CoV-2-negative tests | Qatar’s population | All persons who received a vaccine other than BNT162b2 or mRNA-1273, or individuals who received a fourth vaccine dose (second booster dose) prior to the study SARS-CoV-2 test who received a different mix of vaccines, or tests occurring within 14 days of a second dose or 7 days of a third dose, or SARS-CoV-2 tests conducted because of travel-related requirements. Cases or controls with SARS-CoV-2-positive tests <90 days before the study’s SARS-CoV-2 test were excluded | SARS-CoV-2 reinfection is conventionally defined as a documented infection ≥90 days after an earlier infection | Sex, 10-year age group, nationality, number of coexisting conditions, method of testing, reason for SARS-CoV-2 testing, status of most recent prior infection, and the calendar month of testing |
| Ranzani3 (2022) (53) | Those from the study population who had Covid 19 symptoms, defined by the presence of at least one symptom: fever, sore throat, headache, cough, chills, runny nose, dyspnea, anosmia, and ageusia, and a positive SARS-CoV-2 RT-PCR/antigen test result | Those from the study population who had Covid-19 symptoms as defined by cases, and a negative SARS-CoV 2 RT-PCR /antigen test result | Adults (aged ≥18 years) residing in Brazil, and who underwent SARS-CoV-2 RT-PCR or rapid antigen testing associated with symptomatic illness | Individuals with missing or inconsistent information on age, sex, municipality of residence, and on vaccination and testing status and dates, or RT-PCR/antigen tests that were not collected within 10 days of symptom onset to avoid potentially misclassification, positive or negative RT-PCR/antigen tests with a positive RT-PCR/ antigen test in the previous 90daystocaptureonlyincidentinfections and avoid a second positive test because of prolonged viral shedding, and negative RT-PCR/antigen tests with a positive RT-PCR/antigen test occurring in the following 14 days because of likely false-negative test in the first negative test. All RT-PCR/antigen tests that were obtained after receipt of a primary series of ChAdOx1 nCoV-19, BNT162b2 or Ad26.COV2.S vaccines | Symptomatic Covid-19 includes mild and severe cases. Severe Covid-19 was defined as hospital admission and death with severe acute respiratory infection due to SARS-CoV-2 (positive RT-PCR/Antigen test) | Age, sex, self-reported race, number of chronic comorbidities, previous symptomatic events notified to the surveillance system, municipality of residence, and RTPCR/antigen test sample collection date |
| Roberts (2022) (54) | SARS-CoV-2-positive tests | SARS-CoV-2-negative tests | Adults (aged ≥18 years) who received primary or other health care at MM and had a reverse transcription-polymerase chain reaction (RT-PCR) test for SARS-CoV-2 infection performed or recorded at MM between January 1 and December 31, 2021 | Antigen test results, or partially vaccinated (i.e., had not completed the vaccination primary series or received a mixed sequence of vaccines outside of CDC guidelines) or received unspecified or other vaccine brands (i.e., Astra Zeneca, Sinopharm, Sinovac, or Novavac) | (1) SARS-CoV-2 infection and (2) severe disease or death. SARS-CoV-2 infection was defined as any documented positive result on an RT-PCR test (symptomatic or asymptomatic, non-severe or severe disease), whereas severe disease or death was defined as either hospitalization or intensive care unit (ICU) admission between 7 and +30 days relative to the date of a documented positive SARS-CoV-2 RT-PCR test result or death between 7 and +60 days relative to the date of a positive RT-PCR test | Age, gender, race/ethnicity, Elixhauser score AHRQ, Persons per square mile, neighborhood disadvantage index, past COVID-19 infection, healthcare worker status |
| Rosa (2023) (55) | Positive Reverse transcription polymerase chain reaction (RT-PCR) test for SARS- CoV-2 | Negative Reverse transcription polymerase chain reaction (RT-PCR) test for SARS- CoV-2 | (1) age ≥12 years old; (2) residency in Toledo; (3) seeking care in the public health system of Toledo with symptoms suggestive of COVID- 19 defined as follows: acute respiratory infection symptoms (nasal congestion, rhinorrhea, anosmia, sore throat, hoarseness, new or increased-from-baseline cough, sputum production, dyspnea, wheezing, myalgia) OR diagnosis suggestive of acute respiratory infection symptoms (pneumonia, upper respiratory infection, bronchitis, influenza, cough, asthma, viral respiratory illness, respiratory distress, respiratory failure), and (4) having a nasopharyngeal or nasal sample for SARS-CoV- 2 diagnosis obtained as standard of care | Patients treated with remdesivir in the past 30 days, convalescent plasma treatment in the past 90 days, or monoclonal antibodies for COVID-19 in the past 90 days, patients with unknown vaccination status, and those who refused to provide informed consent | Symptomatic COVID-19, defined by the presence of at least one COVID-19-like symptom as listed in Participants section and confirmed by a positive RT-PCR result for SARS-CoV-2 infection from participants’ nasopharyngeal or nasal swabs | Age group, sex, Charlson comorbidity index, body mass index, current smoking status, and quartiles of time elapsed between the study start and date of RT-PCR sample collection |
| Self (2021) (56) | Admitted to a hospital with COVID-19–like illness (having one or more of the following: fever, cough, shortness of breath, loss of taste, loss of sense of smell, use of respiratory support for the acute illness, or new pulmonary findings on chest imaging consistent with pneumonia) | Test-negative controls (persons with COVID-19–like illness who received negative SARS-CoV-2 RT-PCR test results) and syndrome negative controls (a second control group of persons without COVID-19–like illness who also received negative SARS-CoV-2 RT-PCR test results) | Adults aged ≥18 years who were hospitalized at 21 U.S. hospitals across 18 states during March 11–August 15, 2021 | Patients with immunocompromising conditions | COVID-19 hospitalizations | Admission date, geographic region, age, sex, and race and Hispanic ethnicity |
| Skowronski (2022) (57) | SARS-CoV-2 NAAT-positive | NAAT-negative | Adults ≥18-years-old between epi-weeks 22-47 and assessed for SARS3 CoV-2 by publicly-funded nucleic-acid-amplification-test (NAAT) | Those who had received just one dose of any vaccine | Alpha and Delta variant infection and hospitalization | Age group, sex (men/women), epi-week (categorical) and region |
| Sritipsukho2 (2023) (58) | PUI whose initial or subsequent nasopharyngeal RT–PCR tests were positive | Those with negative initial and all follow-up RT-PCR tests during the 14-day period | Adults ≥18 years old at-risk for COVID-19 who presented TUH or TFH, met the criteria for patients under investigation (PUI) for COVID-19 and received nasopharyngeal RT–PCR test for SARS-CoV-2 at either facility | Those who had prior COVID-19 and/or did not complete the 14-day follow-up period to assess COVID-19 development | SARS-CoV-2 infections | Age, sex, educational level, being healthcare worker, and having any comorbidities at-risk for disease progression |
| Stowe (2022) (59) | Those testing positive by PCR | Those testing negative by PCR | Those aged 18 years and over | For analyses that involved hospitalised controls, any negative tests that led to hospitalisation within 21days of a previous hospital negative test were excluded. Positive and negative tests within 90 days of a previous positive test, or negative tests taken within 21 days after a positive test or a previous negative test that led to a hospitalisation within the previous 21 days | Hospitalization | Sex, index of multiple deprivation, ethnic group, care home residence status, geographic region, period, health and social care worker status, clinical risk group status, clinically extremely vulnerable, severely immunosuppressed, and previously tested positive |
| Suarez (2022) (60) | Respectively positive to a given variant for the analysis by variant | Symptomatic negative individuals | Individuals aged 18 years or over who reported symptoms in the 7days before the time of screening | All individuals with a confirmed SARS-CoV-2 infection at least 60 days prior to the time of screening; individuals with atypical vaccination schemes: those vaccinated with two doses from two different vaccines when one of them was the Janssen vaccine, and those who never received their second dose of a bi-dose vaccine | Symptomatic infections and severe COVID-19 outcomes | Age (ten-year age brackets), sex, area of residence, week of testing and presence or absence of a comorbidity qualifying for prioritization in the vaccination campaign |
| Suphanchaimat (2022) (61) | A Thai national with positive SARS-CoV-2 by PCR test | A Thai national showing negative PCR test for SARS-CoV-2 or professional use Ag-RDT | The whole Thai population | Case (and their matched controls) whose laboratory collection occurred within fourteen days after the last vaccination | SARS-CoV-2 infections | Age, laboratory detection date, and provincial residence of testing sites |
| Surie1 (2022) (62) | Those who received a positive SARS-CoV-2 nucleic acid amplification test (NAAT) or antigen test result within 14 days of illness onset | Those who received a negative SARS-CoV-2 NAAT result | Adults aged ≥18 years admitted for COVID-19–like illness within the IVY Network of 21 hospitals in 18 states | Those who had an immunocompromising condition, or other exclusion criteria: 1) receipt of a non-mRNA vaccine; 2) partial vaccination, including receipt of only 1 mRNA vaccine dose; 3) inability to verify vaccination status; 4) vaccination before CDC recommendations; 5) illness onset >10 days before test date; 6) illness onset >14 days before hospitalization; 7) missing data; and 8) withdrawal from participation | Hospitalization | Date of hospital admission, U.S. Department of Health and Human Services region, age group, sex, and race or ethnicity |
| Tabak (2021) (63) | Test positive cases | Test negative controls | Unique adult persons (18 years or older) who were tested during May 1 through August 7, 2021 at any CVS SARS-CoV-2 test site and reported at least one symptom | Persons with rapid SARS-CoV-2 antigen tests | Symptomatic SARS-CoV-2 infections confirmed by positive polymerase chain reaction tests in US retail locations | Age, region, and calendar month of test |
| Tamada (2023) (64) | People who tested positive | Individuals who tested negative | Adults aged ≥20 years who underwent polymerase chain reaction (PCR) tests or antigen tests in medical institutions as administrative inspections | 1) Vaccination status unknown; 2) comorbidity status was uncertain; 3) previously tested positive before the study period | SARS-CoV-2 infections | Sex, age group, number of comorbidities, residential municipality, and calendar week |
| Tamandjou (2023) (65) | Individuals with at least one SARS-CoV-2 positive RT-PCR test | Those with a negative SARS-CoV 2 PCR test performed within the seven days following the onset of symptoms, and neither preceded nor followed by a positive RT-PCR test during the study period | Individuals aged 50 years and above, with self-reported COVID 19-like symptoms and SARS-CoV-2 RT-PCR tests recorded within the period of June 6, 2021 and February 10, 2022 | Individuals who had received only one vaccine dose as a complete primary vaccination, in presence of a history of documented previous infection, or immunocompromised individuals, or negative antigenic tests without follow-up negative PCR confirmed test and negative PCR tests performed more than seven days from the onsets of symptoms | (1) Symptomatic infection through the TND study, and (2) severe outcomes following a symptomatic infection through the nested cohort study. Two levels of severity of COVID-19 disease were examined: i) any hospitalization, and ii) ICU admissions or in-hospital deaths | Age, sex, type of residence, presence of at least one low or medium-risk comorbidity, and healthcare professional status |
| Tan (2023) (66) | Individuals with a positive PCR | Individuals who tested PCR-negative | Singapore residents aged 18 years without previous SARS-CoV-2 infection who underwent a PCR test at any acute hospital in Singapore | NA | SARS-CoV-2 infections | Sex, age, race, housing type, and calendar week |
| Tartof (2022) (67) | Those with a diagnosis of acute respiratory infection and a KPSC laboratory-confirmed positive SARS-CoV-2 PCR test from a sample collected within 14 days before the initial admission date through to 3 days after the admission | Those with a KPSC laboratory-confirmed negative SARS-CoV-2 PCR test collected within 14 days before a hospital or emergency department admission for acute respiratory infection to 3 days after the admission, and no laboratory-confirmed positive SARS-CoV-2 PCR tests within 90 days before the initial admission | Aged 18 years and older admitted to hospital or an emergency department (without a subsequent hospital admission) with a diagnosis of acute respiratory infection and tested for SARS-CoV-2 via PCR | Individuals with partial vaccination | Hospital and emergency department admissions due to the omicron and delta variants | Age, sex, race/ethnicity, BMI, Charlson comorbidity index, prior SARS-CoV-2 infection, prior influenza vaccination, prior pneumococcal vaccination |
| Tarof2 (2023) (68) | Children testing positive for SARS-CoV-2 | Those who tested negative | Children 5–11 years of age with an ED/UC encounter (without subsequent hospitalization) from November 1, 2021 through September 2, 2022 with a diagnosis of ARI and an RT-PCR test for SARS-CoV-2 | Children with ARI encounters occurring during the 14-day window after dose two, or those who received only a single dose of BNT162b2, >3 doses of BNT162b2, or a COVID-19 vaccine other than BNT162b2, or index < 14 days since vaccination, or unable to classify after whole genome sequencing | COVID-19-associated emergency department or urgent care (ED/UC) encounters | Age, sex, race/ethnicity, prior laboratoryconfirmed SARS-CoV-2 infection, child body mass index category, and pediatric comorbidity index |
| Tarof3 (2022) (69) | Patients with RT-PCR test results positive for SARS-CoV-2 | Patients whose RT-PCR results were negative for SARS-CoV-2 and had not had any positive SARS-CoV-2 test results within 30 days prior to the Ed or UC encounter | Adolescents aged 12 to 17 years with an ED or UC encounter (without subsequent hospitalization) from November 1, 2021, through March 18, 2022, with a diagnosis of acute respiratory infection (ARI)and a reverse transcription-polymerase chain reaction (RT-PCR) nucleic acid amplification test for SARS-CoV-2 from a sample collected within 14 days prior through 3 days after the visit | Participants were excluded if they received only 1 dose of BNT162b2, if they had not yet reached 14 days after receipt of the second dose of BNT162b2, if they had not yet reached 14 days after receipt of the third dose of BNT162b2, if they received a third dose of BNT162b2 less than 5 months after their second dose, or if they received any COVID-19 vaccine other than BNT162b2 | ED and UC encounters related to Delta or Omicron variant SARS-CoV-2 infection | Age, sex, race and ethnicity, body mass index, prior documented positive RT-PCR test result, a pediatric comorbidity index, and encounter date |
| Tartof4 (2022) (70) | Those who tested positive for SARS-CoV-2 | Those who tested negative | KPSC members ≥18 years of age between May 9 and August 26, 2022 who were diagnosed with acute respiratory infection and tested for SARS-CoV-2 on polymerase chain reaction in one of four healthcare settings including outpatient visits (which included virtual appointments), urgent care centers, emergency departments (ED), or the hospital | Immunocompromised individuals yielded similar results, or individuals who received a COVID-19 vaccine other than BNT162b2, who received only one dose of BNT162b2, or received >3 doses | SARS-CoV-2 infections | Week of COVID-19 health-care encounter, age, sex, race or ethnicity, previous SARS-CoV-2 infection, BMI, Charlson score, and history of previous influenza and pneumococcal vaccination, and nirmatrelvir plus ritonavir receipt |
| Tartof5 (2023) (71) | Patients with a hospital or emergency department admission for acute respiratory infection and a positive KPSC laboratory-confirmed SARS-CoV-2 RT-PCR test from a sample collected from 14 days before the initial admission date to 3 days after the admission | Patients with a hospital or emergency department admission for acute respiratory infection and a KPSC laboratory-confirmed negative SARS-CoV-2 RT-PCR test collected from 14 days before admission to 3 days after admission and no positive COVID-19 tests within 90 days of admission | Adult (aged ≥18 years) members of Kaiser Permanente Southern California (KPSC) | Members younger than 18 years, or individuals who had received partial (ie, one dose) or heterologous vaccination | Emergency department or hospital admission | The month of admission, age, sex, race and ethnicity, body-mass index, Charlson Comorbidity Index, previous influenza vaccination, previous pneumococcal vaccination, and previous SARS-CoV-2 infection |
| Tenforde3 (2022) (72) | Had COVID-19-like illness (CLI) and tested positive 4 for SAR-CoV-2 by molecular or antigen test within 10 days of illness onset | Test-negative control (patients hospitalized with CLI who tested negative for SARS-CoV-2 by RT-PCR); syndrome-negative control (patients hospitalized without CLI who tested negative for SARS-CoV-2 by RT-PCR) | Adults (≥18 years) hospitalized at 21 hospitals in 18 states March 11-December 15, 2021 | Patients who received 1 or more doses of a mRNA vaccine but did not meet study criteria for full vaccination, who received mixed vaccine products, or who received a non-mRNA vaccine, as well as patients who received more than two doses of a mRNA vaccine with the third dose received ≥7 days before illness onset | Hospitalization | Calendar date of admission (in biweekly intervals), age, sex, and race and ethnicity, presence of underlying chronic conditions, immunocompromised status, and US Health and Human Services region of the admitting hospital |
| Thompson (2021) (73) | A person with at least one ICD code that was consistent with Covid-19–like illness and at least one positive SARS-CoV-2 test result | Persons with at least one ICD code for Covid-19–like illness and only negative SARSCoV-2 test results | Adults (≥50 years of age) with Covid-19–like illness who underwent molecular testing for severe acute respiratory syndrome coronavirus 2 (SARS-CoV-2) | Patients tested using only a non-molecular assay; adults hospitalized prior to age-specific inclusion dates; patients receiving dose-1 <14 days prior to index hospitalization date | Laboratory-confirmed SARS-CoV-2 CLI-associated medical event, which is based on clinical testing and clinical diagnosis | Age, geographic region, calendar time, and local virus circulation |
| Thompson2 (2022) (74) | Those who with acute laboratory-confirmed COVID-19 | Those who without acute COVID-19 | Those among adults aged ≥18 years with a COVID-19-like illness diagnosis who had received molecular testing (primarily reverse transcription-polymerase chain reaction assay) for SARS-CoV-2 (the virus that causes COVID-19) within 14 days before or 72 hours after the admission or encounter | Recipients of Ad26.COV2 (Janssen [Johnson & Johnson]), 1 or >3 doses of an mRNA vaccine, and those for whom 1-13 days had elapsed since receipt of any dose were excluded | Emergency Department and Urgent Care Encounters and Hospitalizations | Age, geographic region, calendar time, and local virus circulation and weighted for inverse propensity to be vaccinated or unvaccinated (calculated separately for each VE estimate |
| Tseng (2022) (75) | Individuals testing positive for SARS-CoV-2 | Individuals testing negative | Individuals who tested with specimens collected between 6 December 2021 and 31 December 2021, were aged ≥18 years and had ≥12 months of KPSC membership before the specimen collection date | Received a COVID-19 vaccine other than mRNA-1273, any dose of mRNA-1273; a positive SARS-CoV-2 test or COVID-19 diagnosis code ≤90 days before the specimen collection date | Infection, COVID-19 hospitalization included hospitalization with a SARS-CoV-2-positive test or hospitalization ≤7 days after a SARS-CoV-2-positive test | History of SARS-CoV-2 molecular test, preventive care, Charlson comorbidity score, obesity, immunocompromised status and history of COVID-19 |
| van Ewijk (2023) (76) | Persons testing SARS-CoV-2 positive | Persons testing SARS-CoV-2 negative | Individuals who were 18 years and older and completed the questionnaire before receiving test result, did not reside at a care facility and had not previously participated in the study | People who were partially vaccinated, received a booster (second vaccination in case of Janssen, third vaccination in case of the other vaccines) or third vaccination and those with heterologous vaccination schemes, with missing data regarding vaccination status, with onset of symptoms > 10 days before getting tested, people who reported a previous laboratory-confirmed SARS-CoV-2 infection, those with inconclusive test results and people who went to the test facility for confirmation of a positive self-administered LFAT to minimise recall bias | SARS-CoV-2 infections | Age, sex, calendar week, education level, comorbidities, household size, number of close contacts inside and outside, face mask wearing habits, visiting busy locations inside and outside, contact with a SARS-CoV-2-positive person |

Table S4. Summary of number of studies and estimates by key factors

|  | Infection | | Severe disease | |
| --- | --- | --- | --- | --- |
|  | Number of studies | Number of estimates | Number of studies | Number of estimates |
| Handling of participants with prior infection | | | | |
| Included | 44 | 522 | 39 | 343 |
| Excluded | 25 | 402 | 12 | 152 |
| Enrolment criteria | | | | |
| Clinical | 41 | 579 | 32 | 292 |
| Non-clinical | 22 | 345 | 17 | 203 |
| Vaccine type |  |  |  |  |
| mRNA | 56 | 812 | 40 | 389 |
| Adenovirus vector | 10 | 78 | 10 | 62 |
| Inactivated | 8 | 34 | 9 | 44 |
| Predominant circulating virus | | | | |
| Pre-Delta and Delta | 16 | 179 | 19 | 177 |
| Late-Delta | 38 | 306 | 24 | 171 |
| Omicron (BA.1/BA.2) | 35 | 320 | 22 | 120 |
| Omicron (BA.4/BA.5) and afterwards | 9 | 119 | 8 | 27 |

Table S5. Number of estimates available disaggregated by inclusion of participants with prior infection

|  | Included participants with prior infection | | | | Excluded participants with prior infection | | | |
| --- | --- | --- | --- | --- | --- | --- | --- | --- |
|  | Infection | | Severe disease | | Infection | | Severe disease | |
|  | Number of studies | Number of estimates | Number of studies | Number of estimates | Number of studies | Number of estimates | Number of studies | Number of estimates |
| Handling of participants with prior infection |  |  |  |  |  |  |  |  |
| Included | 44 | 522 | 39 | 343 |  |  |  |  |
| Excluded |  |  |  |  | 25 | 402 | 12 | 152 |
| Enrolment criteria |  |  |  |  |  |  |  |  |
| Clinical | 30 | 261 | 25 | 191 | 16 | 318 | 8 | 101 |
| Non-clinical | 14 | 261 | 14 | 152 | 9 | 84 | 4 | 51 |
| Vaccine type |  |  |  |  |  |  |  |  |
| mRNA | 37 | 452 | 30 | 251 | 24 | 360 | 12 | 138 |
| Adenovirus vector | 5 | 41 | 8 | 49 | 7 | 37 | 3 | 13 |
| Inactivated | 6 | 29 | 8 | 43 | 2 | 5 | 1 | 1 |
| Predominant circulating virus |  |  |  |  |  |  |  |  |
| Pre-Delta and Delta | 11 | 110 | 15 | 118 | 6 | 69 | 5 | 59 |
| Late-Delta | 27 | 213 | 18 | 133 | 13 | 93 | 7 | 38 |
| Omicron (BA.1/BA.2) | 24 | 157 | 17 | 72 | 15 | 163 | 5 | 48 |
| Omicron (BA.4/BA.5) and afterwards | 8 | 42 | 7 | 20 | 2 | 77 | 2 | 7 |

Table S6. Relationship between government response index and estimates of risk ratios against infection or severe disease.

| Endpoint | Infection | Infection | Infection | Severe disease | Severe disease | Severe disease |
| --- | --- | --- | --- | --- | --- | --- |
| Index | Stringency index | Containment health index | Government response index | Stringency index | Containment health index | Government response index |
| *Model 1: Model adjusted for vaccine type, circulating virus and recruitment criteria + day since vaccination + study including participants with COVID infection history* | | | | | | |
| Prior infection | 1.04 (0.98, 1.12) | 1.04 (0.98, 1.12) | 1.04 (0.98, 1.12) | 1.39 (1.19, 1.62) | 1.39 (1.19, 1.62) | 1.39 (1.19, 1.62) |
| Vaccine type | | | | | | |
| mRNA vaccines | REF | REF | REF | REF | REF | REF |
| Adenovirus vector vaccines | 1.51 (1.34, 1.69) | 1.49 (1.32, 1.67) | 1.50 (1.33, 1.68) | 2.07 (1.66, 2.58) | 2.04 (1.63, 2.56) | 2.07 (1.65, 2.59) |
| inactivated virus vaccines | 2.40 (2.05, 2.82) | 2.24 (1.90, 2.63) | 2.23 (1.89, 2.62) | 3.58 (2.80, 4.58) | 3.46 (2.67, 4.49) | 3.54 (2.72, 4.61) |
| Circulating variants | | | | | | |
| pre-Delta and Delta | REF | REF | REF | REF | REF | REF |
| late delta | 0.95 (0.86, 1.04) | 0.98 (0.89, 1.07) | 0.97 (0.89, 1.07) | 0.91 (0.75, 1.09) | 1.00 (0.83, 1.21) | 1.03 (0.85, 1.24) |
| Omicron (BA.1/BA.2) | 2.74 (2.46, 3.05) | 2.96 (2.66, 3.29) | 2.99 (2.69, 3.32) | 3.38 (2.64, 4.32) | 4.12 (3.24, 5.24) | 4.36 (3.42, 5.57) |
| Omicron (BA.4/BA.5) and afterwards | 2.43 (2.09, 2.82) | 2.77 (2.39, 3.22) | 2.76 (2.38, 3.21) | 4.28 (2.76, 6.63) | 6.05 (3.88, 9.44) | 6.85 (4.43, 10.59) |
| Timing since vaccination | | | | | | |
| 0-30 | REF | REF | REF | REF | REF | REF |
| 31-60 | 0.97 (0.84, 1.13) | 0.95 (0.81, 1.10) | 0.95 (0.82, 1.10) | 0.73 (0.51, 1.04) | 0.71 (0.49, 1.01) | 0.71 (0.49, 1.01) |
| 61-90 | 1.34 (1.15, 1.57) | 1.33 (1.14, 1.56) | 1.33 (1.14, 1.56) | 1.13 (0.79, 1.62) | 1.14 (0.79, 1.64) | 1.15 (0.79, 1.66) |
| 91-120 | 1.33 (1.15, 1.54) | 1.30 (1.12, 1.51) | 1.30 (1.12, 1.51) | 0.76 (0.54, 1.06) | 0.74 (0.53, 1.04) | 0.74 (0.53, 1.05) |
| 121-180 | 1.66 (1.43, 1.93) | 1.65 (1.41, 1.92) | 1.64 (1.41, 1.92) | 1.13 (0.80, 1.60) | 1.13 (0.79, 1.60) | 1.13 (0.79, 1.61) |
| 181+ | 2.04 (1.79, 2.33) | 2.01 (1.76, 2.30) | 2.01 (1.76, 2.31) | 1.05 (0.77, 1.45) | 1.03 (0.75, 1.42) | 1.03 (0.74, 1.42) |
| Model 1 + addition variables below as three separate models | | | | | | |
| Stringency index | 0.84 (0.81, 0.87) | NA | NA | 0.84 (0.77, 0.92) | NA | NA |
| Containment health index | NA | 0.83 (0.79, 0.88) | NA | NA | 0.90 (0.79, 1.02) | NA |
| Government response index | NA | NA | 0.85 (0.81, 0.89) | NA | NA | 0.95 (0.84, 1.07) |

Table S7. Relationship between government response index and estimates of risk ratios against infection or severe disease, excluding estimates from studies rated that serious risk of bias

| Endpoint | Infection | Infection | Infection | Severe disease | Severe disease | Severe disease |
| --- | --- | --- | --- | --- | --- | --- |
| Index | Stringency index | Containment health index | Government response index | Stringency index | Containment health index | Government response index |
| *Model1: Model adjusted for vaccine type, circulating virus and recruitment criteria + day since vaccination + study including participants with COVID infection history* | | | | | | |
| Prior infection | 1.01 (0.93, 1.10) | 1.01 (0.93, 1.10) | 1.01 (0.93, 1.10) | 1.38 (1.18, 1.62) | 1.38 (1.18, 1.62) | 1.38 (1.18, 1.62) |
| Vaccine type | | | | | | |
| mRNA vaccines | REF | REF | REF | REF | REF | REF |
| Adenovirus vector vaccines | 1.52 (1.34, 1.72) | 1.50 (1.32, 1.71) | 1.51 (1.33, 1.72) | 2.05 (1.65, 2.56) | 2.02 (1.61, 2.54) | 2.05 (1.64, 2.57) |
| inactivated virus vaccines | 2.48 (2.09, 2.94) | 2.33 (1.96, 2.77) | 2.32 (1.95, 2.76) | 3.54 (2.77, 4.53) | 3.43 (2.64, 4.45) | 3.50 (2.69, 4.57) |
| Circulating variants | | | | | | |
| pre-Delta and Delta | REF | REF | REF | REF | REF | REF |
| late delta | 0.99 (0.89, 1.09) | 1.02 (0.93, 1.13) | 1.03 (0.93, 1.14) | 0.89 (0.74, 1.08) | 0.99 (0.82, 1.20) | 1.02 (0.84, 1.23) |
| Omicron (BA.1/BA.2) | 2.75 (2.44, 3.09) | 2.94 (2.62, 3.31) | 3.04 (2.71, 3.41) | 3.33 (2.60, 4.26) | 4.08 (3.21, 5.20) | 4.32 (3.39, 5.51) |
| Omicron (BA.4/BA.5) and afterwards | 2.58 (2.08, 3.21) | 3.01 (2.44, 3.73) | 3.14 (2.54, 3.88) | 4.20 (2.71, 6.53) | 5.99 (3.84, 9.36) | 6.78 (4.39, 10.48) |
| Timing since vaccination | | | | | | |
| 0-30 | REF | REF | REF | REF | REF | REF |
| 31-60 | 0.90 (0.76, 1.07) | 0.87 (0.73, 1.04) | 0.87 (0.73, 1.04) | 0.71 (0.50, 1.02) | 0.69 (0.48, 0.998) | 0.69 (0.48, 1.001) |
| 61-90 | 1.27 (1.05, 1.52) | 1.26 (1.04, 1.52) | 1.25 (1.03, 1.50) | 1.11 (0.77, 1.60) | 1.13 (0.78, 1.63) | 1.13 (0.78, 1.64) |
| 91-120 | 1.22 (1.02, 1.45) | 1.19 (0.999, 1.42) | 1.18 (0.99, 1.41) | 0.75 (0.54, 1.05) | 0.74 (0.52, 1.04) | 0.74 (0.52, 1.04) |
| 121-180 | 1.55 (1.30, 1.86) | 1.54 (1.28, 1.84) | 1.52 (1.27, 1.83) | 1.11 (0.78, 1.57) | 1.10 (0.77, 1.58) | 1.11 (0.77, 1.58) |
| 181+ | 1.85 (1.58, 2.17) | 1.81 (1.54, 2.13) | 1.81 (1.53, 2.13) | 1.04 (0.75, 1.43) | 1.01 (0.73, 1.41) | 1.01 (0.73, 1.41) |
| Model 1 + addition variables below as three separate models | | | | | | |
| Stringency index | 0.85 (0.81, 0.89) | NA | NA | 0.84 (0.77, 0.92) | NA | NA |
| Containment health index | NA | 0.84 (0.79, 0.89) | NA | NA | 0.90 (0.79, 1.02) | NA |
| Government response index | NA | NA | 0.87 (0.82, 0.92) | NA | NA | 0.95 (0.84, 1.07) |

Table S8. Relationship between government response index and estimates of risk ratios against infection or severe disease, excluding estimates from studies not using clinical case definition or excluding participants with prior infections.

|  | Endpoint | Infection | Infection | Infection | Severe disease | Severe disease | Severe disease |
| --- | --- | --- | --- | --- | --- | --- | --- |
|  | Index | Stringency index | Containment health index | Government response index | Stringency index | Containment health index | Government response index |
|  | *Model 1: Model adjusted for vaccine type, circulating virus and recruitment criteria + day since vaccination + study including participants with COVID infection history* | | | | | | |
|  | Prior infection | 0.91 (0.84, 0.99) | 0.91 (0.84, 0.99) | 0.91 (0.84, 0.99) | 1.20 (1.004, 1.44) | 1.20 (1.004, 1.44) | 1.20 (1.004, 1.44) |
|  | Vaccine type | | | | | | |
|  | mRNA vaccines | REF | REF | REF | REF | REF | REF |
|  | Adenovirus vector vaccines | 1.60 (1.42, 1.81) | 1.58 (1.40, 1.79) | 1.61 (1.42, 1.82) | 2.53 (2.03, 3.16) | 2.51 (2.01, 3.15) | 2.49 (1.99, 3.12) |
|  | inactivated virus vaccines | 2.38 (2.03, 2.80) | 2.17 (1.84, 2.56) | 2.11 (1.79, 2.49) | 3.43 (2.72, 4.32) | 3.33 (2.61, 4.24) | 3.18 (2.48, 4.08) |
|  | Circulating variants | | | | | | |
|  | pre-Delta and Delta | REF | REF | REF | REF | REF | REF |
|  | late delta | 0.89 (0.81, 0.98) | 0.89 (0.81, 0.99) | 0.87 (0.78, 0.96) | 0.86 (0.71, 1.04) | 0.89 (0.73, 1.07) | 0.85 (0.70, 1.03) |
|  | Omicron (BA.1/BA.2) | 2.72 (2.43, 3.05) | 2.80 (2.49, 3.15) | 2.69 (2.40, 3.02) | 3.50 (2.75, 4.45) | 3.71 (2.92, 4.72) | 3.45 (2.68, 4.44) |
|  | Omicron (BA.4/BA.5) and afterwards | 2.45 (2.09, 2.87) | 2.64 (2.25, 3.10) | 2.44 (2.08, 2.86) | 5.54 (3.62, 8.47) | 6.18 (4.03, 9.47) | 5.55 (3.62, 8.51) |
|  | Timing since vaccination | | | | | | |
|  | 0-30 | REF | REF | REF | REF | REF | REF |
|  | 31-60 | 1.02 (0.88, 1.19) | 1.01 (0.86, 1.18) | 1.02 (0.88, 1.19) | 0.79 (0.55, 1.14) | 0.79 (0.55, 1.14) | 0.79 (0.55, 1.13) |
|  | 61-90 | 1.38 (1.18, 1.62) | 1.39 (1.18, 1.63) | 1.40 (1.20, 1.64) | 1.26 (0.88, 1.80) | 1.27 (0.89, 1.82) | 1.26 (0.88, 1.80) |
|  | 91-120 | 1.35 (1.16, 1.57) | 1.33 (1.14, 1.56) | 1.35 (1.16, 1.57) | 0.88 (0.63, 1.23) | 0.88 (0.63, 1.24) | 0.88 (0.63, 1.23) |
|  | 121-180 | 1.68 (1.44, 1.96) | 1.68 (1.44, 1.97) | 1.69 (1.45, 1.97) | 1.11 (0.78, 1.57) | 1.11 (0.78, 1.58) | 1.10 (0.78, 1.57) |
|  | 181+ | 1.85 (1.58, 2.17) | 1.81 (1.54, 2.13) | 1.81 (1.53, 2.13) | 1.04 (0.75, 1.43) | 1.01 (0.73, 1.41) | 1.01 (0.73, 1.41) |
| Model 1 + addition variables below as three separate models | | | | | | | |
| Stringency index | | 0.83 (0.80, 0.87) | NA | NA | 0.92 (0.85, 1.005) | NA | NA |
| Containment health index | | NA | 0.80 (0.76, 0.85) | NA | NA | 0.93 (0.83, 1.05) | NA |
| Government response index | | NA | NA | 0.80 (0.76, 0.84) | NA | NA | 0.89 (0.79, 1.01) |

Table S9. Variables in extraction form

| variable | description |
| --- | --- |
| study id | Identity number of the study |
| author | first author of the study |
| year | publish year |
| paper | published paper |
| pmid | PubMed ID of the study |
| study type | general type of the study |
| study | specific type of study |
| study_period_start | the start date of the study period |
| study_period_end | the end date of the study period |
| vaccine | vaccine type used |
| historyofCOVID | Method of controlling covid infection history, included(0): included those with covid history, excluded(1): excluded those with covid history |
| dose_number | total dose number of the vaccine |
| dose | dose number |
| timing_of_dose_days | the time after vaccination |
| outcome | endpoint of the study |
| country | study location |
| population | study population |
| VE | vaccine effectiveness |
| LCI | the lower bound of 95% confidence interval for vaccine effectiveness |
| UCI | the upper bound of 95% confidence interval for vaccine effectiveness |
| no of case(total) | total number of cases |
| no of case(vac) | total number of vaccinated cases |
| no of case(unvac) | total number of unvaccinated cases |
| no of control(total) | total number of control |
| no of control(vac) | total number of vaccinated control |
| no of control(unvac) | total number of unvaccinated control |
| variant | predominant circulating virus during the study period |
| overall | for overall/individual vaccine estimate, NA:that study reported either overall vaccine or individual vaccine estimate; 1:reported both overall vaccine and individual vaccine estimate, and the corresponding row is for overall vaccine; 0:reported both overall vaccine and individual vaccine estimate, and the corresponding row is for individual vaccine |
| adjust for | adjustment of the estimate |
| adjust | the estimate provided is adjusted or not, 1:with adjustment, 0: without adjustment |
| by_clinial | enrolment critera, 1: clinical, 0: non-clinical |
| rob | risk of bias |
| prop_prior inf | proportion of included participants with prior infection |
| method_prior infection | methods for determining prior infection |
| methods_vaccination status | methods for determining vaccination status |
| cases definition | definition of cases |
| controls definition | definition of controls |
| periods_diagnosis tests | periods of diagnosis tests |
| types_diagnosis tests | types of diagnosis tests |
| timing since vaccination included | Detail of included days since vaccination |
| outcomes definition | definition of outcomes |
| inclusion criteria_study population | detailed inclusion criteria of study population |
| exclusion criteria_study population | detailed exclusion criteria of study population |
| ave_stringencyindex | average of stringency index |
| min_stringencyindex | minimum of stringency index |
| max_stringencyindex | maximum stringency index |
| ave_containmenthealthindex | average of containment health index |
| min_containmenthealthindex | minimum of containment health index |
| max_containmenthealthindex | maximum of containment health index |
| ave_governmentresponseindex | average of government response index |
| min_governmentresponseindex | minimum of government response index |
| max_governmentresponseindex | maximum of government response index |
